# Development and validation of a machine learning model for community-based tuberculosis screening among persons aged ≥ 15 years in South Africa and Zambia

**DOI:** 10.64898/2026.03.30.26349632

**Authors:** Alexandra J. Zimmer, Kindie Fentahun Muchie, Henry Loharja, Lisa Koeppel, Helen Ayles, Maria del Mar Castro, Evangelia Christodoulou, Greg J. Fox, Mary Gaeddert, Yohhei Hamada, Chris Isaacs, Nathan Kapata, Pascalina Chanda-Kapata, Kasra Karimi, Nkatya Kasese, Andrew D. Kerkhoff, Irwin Law, Lena Maier-Hein, Florian M. Marx, Minyoi M. Maimbolwa, Sizulu Moyo, Thuli Mthiyane, Monde Muyoyeta, Joacim Rocklöv, Ab Schaap, Seda Yerlikaya, Michael Opata, Claudia M. Denkinger

**Author notes:** Contributed equally (Alexandra J. Zimmer, Kindie Fentahun Muchie, Henry Loharja).

## Abstract

**Introduction:** Current tuberculosis (TB) screening tools, such as the WHO four-symptom screen (W4SS), lack sufficient sensitivity and specificity for effective community-based active case finding, contributing to both missed diagnoses and unnecessary diagnostic evaluations. This study aimed to develop and validate a machine learning (ML) model to improve TB risk prediction among persons aged ≥15 years in community settings of Zambia and South Africa.

**Methods:** A large, harmonized dataset was created from four community-based TB prevalence surveys in South Africa and Zambia (N=169,813), restricted to individuals not under treatment at the time of survey. A binary reference outcome was defined based on available microbiological and radiographic data, grouping individuals as either ‘Possible TB’ or ‘Unlikely TB’. An XGBoost model was trained on 80% (N=135,854) of the data using demographic, clinical, and socio-economic variables, and model interpretability was assessed using SHapley Additive exPlanations (SHAP) values. Internal validation was performed using a 20% hold-out test set (N=33,959). Model performance was assessed using discrimination, calibration, and clinical utility measures compared to the W4SS and against WHO’s 2025 Target Product Profile (TPP) for a tool in a two-step screening algorithm.

**Results:** Overall, 16,413 (9.7%) of individuals were labelled as ‘Possible TB’. On the test set, the XGBoost model yielded an area under the curve (AUC) of 79.7% (95% CI: 78.7, 80.7), outperforming the W4SS (AUC 57.0%; 95% CI: 56.1, 57.8). The XGBoost model achieved 81.5% sensitivity (95% CI: 77.6, 84.9) at a 60% specificity threshold. This exceeded the W4SS, which achieved only 38.2% sensitivity (95% CI: 36.5, 39.9) on the same dataset. SHAP analysis identified age, previous TB treatment, times treated for TB and unemployment as the primary contributors to risk.

**Conclusion:** The ML XGBoost model shows promise as a screening tool to support community-based active case finding activities prior to diagnostic testing. However, as performance remained below TPP targets, and adding variables, e.g. on geolocation, could be considered.

## Introduction

Tuberculosis (TB) remains the world’s leading infectious cause of death, claiming an estimated 1.23 million lives in 2024.^1^ Annually, approximately 30% (2.9 million) of TB-positive individuals go undetected, driving ongoing transmission and mortality.^1,2^ This diagnostic gap stems from reliance on passive case finding, a strategy where individuals self-present to healthcare services.^3^ However, this approach is often hindered by structural barriers, preventing many from obtaining care in a timely manner.^4^ To reach the “missing millions,” the WHO advocates for active case finding (ACF) in high-risk populations, including communities with a prevalence exceeding 0.5%.^2^

Effective ACF is challenged by the suboptimal accuracy and/or accessibility of current screening tools.^5^ Because an estimated 50% of individuals with active TB are asymptomatic,^6,7^ the WHO four-symptom screen (W4SS), which focuses on the presence of cough, fever, weight loss, or night sweats, has poor sensitivity (51-89%).^2,8,9^ While the WHO formally recommends the W4SS as a rule-out tool for people living with HIV (PLWHIV),^9^ the absence of other accessible tools has led to its widespread adoption for screening the general population. Symptom screening also lacks specificity (28–71%), requiring further testing to identify the few individuals with TB, driving costs and limiting scale-up.^9,10^ While more accurate screening tools like chest X-rays (CXR) and rapid molecular tests exist, their use is often restricted to centralized facilities due to high costs and infrastructure requirements.^11^ Consequently, there is a need for more accessible screening tools that can identify high-risk individuals at all levels of care. The 2025 WHO Target Product Profiles (TPP) set minimum benchmarks of 90% sensitivity and 60% specificity for an initial screening test in a multi-step approach where a second screening tool (such as computer aided detection-analyzed CXR) follows to optimize specificity before confirmatory testing.^12^

Advances in artificial intelligence and machine learning (ML) offer a promising avenue to meet this need by leveraging readily available clinical and epidemiological data for TB risk prediction.^13,14^ Combined with a smartphone application, this could be made accessible across different levels of care. However, existing models have yielded limited impact as most models rely on a narrow set of input variables or focus on specific high-risk groups (e.g., PLWHIV, household contacts).^15–18^ Further, no existing prediction model has progressed toward integrating in an accessible screening tool. This study aims to develop and validate a ML prediction model to generate personalized TB risk scores for individuals aged ≥15 years in community settings.

## Methods

### Study design and reporting

This study is a secondary data analysis reported in accordance with the Transparent Reporting of a multivariable prediction model of Individual Prognosis Or Diagnosis for Artificial Intelligence (TRIPOD+AI) guidelines (**Table S1**).^19^

### Datasets and population

Four national and community-based TB prevalence survey datasets from South Africa and Zambia were analyzed: the 2017-2019 South African survey,^6^ the 2013-2014 Zambian survey,^20^ the 2019-2021 TREATS TB survey,^21^ and the 2010 ZAMSTAR survey (**Table S2**).^22^ Eligibility for microbiological testing varied across these studies; the Zambia, South Africa, and TREATS surveys considered symptoms and/or CXR abnormalities (using a CAD4TB threshold of ≥50 in TREATS), while ZAMSTAR performed universal microbiological testing irrespective of symptoms and did not utilize CXR.

For this analysis, the study population was restricted to participants aged ≥15 years not receiving TB treatment during the surveys. Individuals lacking microbiological diagnoses or CXR results were excluded because their outcome status could not be established for model training.

### Outcome definition for model training

A binary outcome (“Possible TB” and “Unlikely TB”) was defined using composite reference standard. Individuals were classified as “Possible TB” if they tested positive on any available microbiological (culture or molecular test) or radiographic assessment (CXR abnormality). Conversely, participants were classified as “Unlikely TB” if they tested negative on all performed assessments within their respective surveys. Self-reported symptoms were excluded from this composite outcome to prevent incorporation bias and ensure they served as independent predictors (**Table S3**).

### Input variables for model development

Inputs comprised 27 variables collected in the prevalence surveys that are easily measured in community-based settings and known to be associated with TB, including demographics, socio-economic indicators, health behavior, self-reported symptoms and duration, TB contact and history, and HIV status. Variables present in at least two data sources with less than 20% missing values were retained and harmonized (**Table S4, Table S5**).

### Data partitioning, sample size, and missing data imputation

The harmonized dataset was split into training (N=135,854; 80%) and evaluation (N=33,959; 20%) sets. The split was stratified by study and outcome to maintain comparable distributions between the training and evaluation sets (**Table S6**). Sample size was fixed based on the data from the prevalence surveys, and adequacy was confirmed using the pmsampsize package in R, based on the Riley et al. criteria for minimum sample size for clinical prediction models.^23,24^ Assuming a 0.1 outcome proportion, 42 candidate parameters (degrees of freedom), a 0.90 shrinkage factor, and 75% area under the curve (AUC), the minimum required sample size was 4,854 participants, well below the training set.

Missing data for the predictors were determined to be missing at random (**Figure S1**) and thus handled using multiple imputation by chained equations (MICE). The mice R package generated 20 imputed datasets separately for the training and test subsets.^25^ Exploratory analyses confirmed that variable distributions remained stable across imputations as compared to the original data (**Figure S2, Figure S3).**

### Model training and evaluation

An XGBoost ensemble model was built in Python (version 3.11.5). Hyperparameters (**Table S7**) were optimized using a Bayesian search cross-validation across five splits of the training set, maximizing on balanced accuracy. The optimal hyperparameter combination identified by this search was used to train the final model. To address class imbalance, inverse proportional weighting was applied during the training process. Separate XGBoost models were trained on each of the 20 imputed datasets and combined into an ensemble model.

Before ensembling, each of the 20 XGBoost models was evaluated on a corresponding test set. Performance was assessed across three clinically meaningful thresholds: 1) the Youden index (maximizing sensitivity and specificity), 2) 90% sensitivity (WHO TPP benchmark for two-step screening), and 3) 60% specificity (WHO TPP benchmark for two-step screening).^12^ Because TPP targets are specifically designed for microbiologically confirmed TB, they were utilized here as heuristic benchmarks for our broader composite outcome. Scores meeting or exceeding the threshold classified individuals as “Possible TB,” while lower scores indicated “Unlikely TB.” Predictive performance was measured using the AUC, sensitivity and specificity, positive (PPV) and negative predictive values (NPV), Brier score, calibration slope, and calibration intercept (**Note S1**). To account for imputation uncertainty, 95% confidence intervals (CIs) for all metrics were computed using Rubin’s combining rules.^26^ This approach incorporated both within-imputation variance (estimated utilizing 1,000 bootstrap resamples per dataset) and between-imputation variance (capturing the variability across the 20 imputed datasets).

### Model interpretability

Model interpretability was assessed using SHapley Additive Explanations (SHAP) values, which quantify the average marginal contribution of each input variable toward a specific prediction relative to a baseline value.^27^ SHAP values were averaged across the 20 models. Global feature importance, determined by calculating the mean absolute SHAP value for each feature across all observations, was visualized alongside the distribution and direction of feature effects using SHAP violin plots. These plots explain the model’s predictions on two levels: how specific predictors impact a single patient’s risk, and how those factors rank in importance across the entire cohort. Bridging these individual and population-level insights verifies that the model’s internal logic aligns with established domain knowledge.

### Comparison with W4SS sub-analysis

To evaluate the model’s performance in relation to existing TB screening methods, the XGBoost model’s predictions were compared to the W4SS on the test set. Individuals presenting with any of the four W4SS symptoms (cough, fever, night sweats, or weight loss) were classified as “Possible TB.” Predictive performance between the W4SS and the XGBoost model was then compared using the AUC, sensitivity and specificity, and PPV and NPV.

### Decision curve analysis for clinical utility

Clinical utility was evaluated and compared to the W4SS using decision curve analysis (DCA) conducted with the rmda R package.^28,29^ DCA quantifies the net benefit of guiding clinical decisions across a range of threshold probabilities (p_t_), representing the minimum predicted risk prompting intervention. This approach defines the acceptable clinical trade-off between false-positive interventions and identifying true-positive cases (**Note S2**). The model’s net benefit was benchmarked against a “Test All” strategy (referring all individuals for confirmatory testing) and a “Test None” strategy (net benefit of zero). Furthermore, clinical utility was assessed via subgroup analyses for sex, HIV status, prior TB treatment, and W4SS symptom presence, utilizing covariate information from the first imputation.

### Ethics

Ethical approval for this secondary data analysis was granted by the Ethics Committee of the Medical Faculty of Heidelberg University (Reference number: S-245/2022).

## Results

### Participant characteristics

A total of 354,834 records were enumerated across the four surveys, yielding 181,437 individuals were included in the final surveys. Of these, 169,813 (93.6%) were included for model training and validation (**Figure 1**).

**Figure 1.**
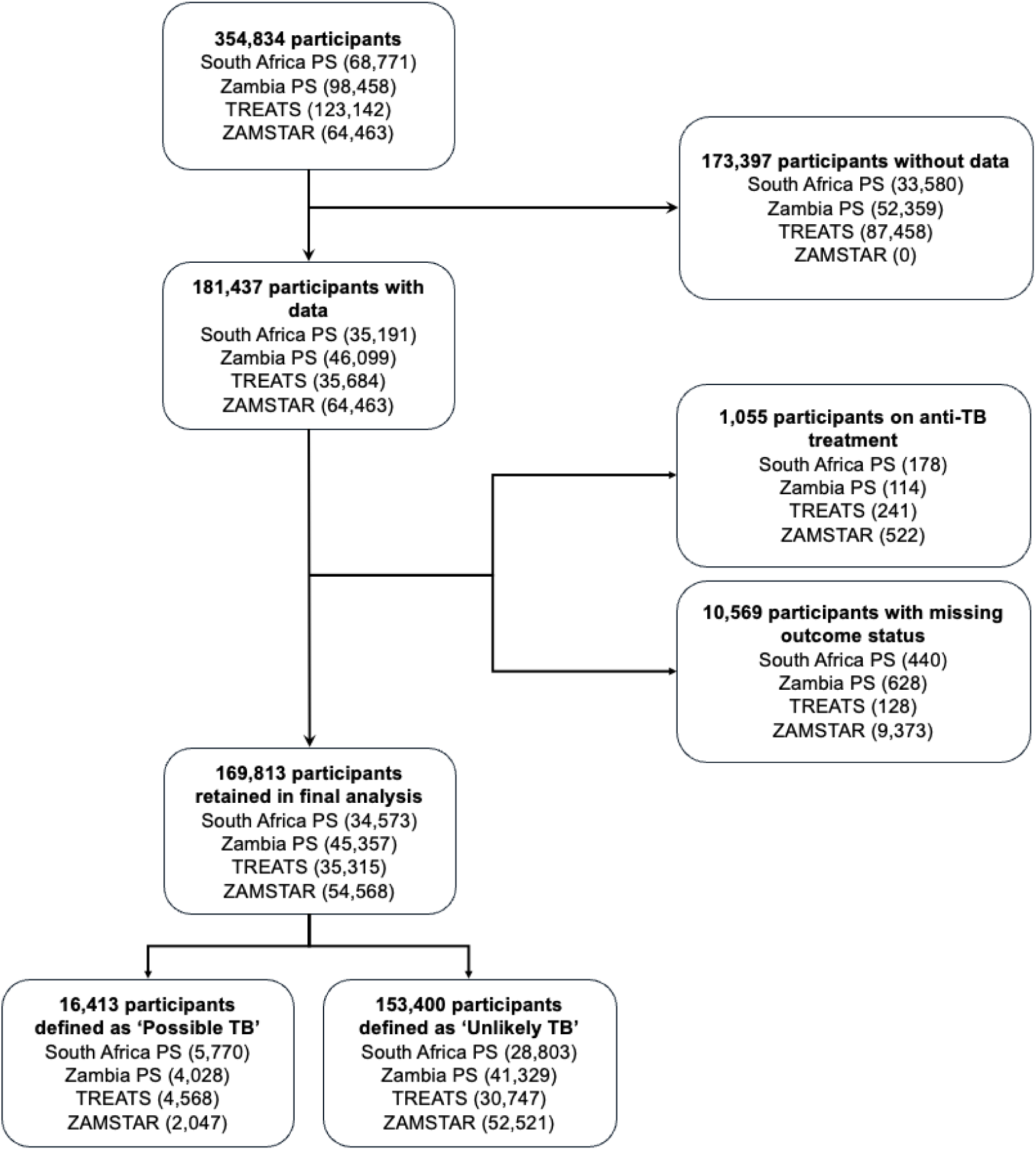
Flow diagram of study participant selection. The diagram details the inclusion and exclusion process across the four data sources. “Records without data” refers to individuals who were enumerated in the initial dataset but for whom no survey data was collected. Numbers in brackets represent the number of records. PS, prevalence survey; TB, tuberculosis; TREATS TB, Tuberculosis Reduction through Expanded Antiretroviral Treatment and Screening for Active Tuberculosis; ZAMSTAR, Zambia, South Africa Tuberculosis and AIDS Reduction. Check details of outcome classification under the respective subheading in methods.

**Table 1** summarizes key participant characteristics from the unimputed datasets. The harmonized dataset (N=169,813) featured a median age of 31 years (IQR: 22-45), with a predominantly female (62.2%) population and 11.2% of PLWHIV. Most participants (70.3%) reported no prior TB treatment, 6.7% were previously treated, and 23.0% had missing treatment data (primarily from the Zambia survey). **Table S3** details the distributions of all variables. Ultimately, 153,400 (90.3%) individuals were classified as “Unlikely TB” and 16,413 (9.7%) as “Possible TB” (**Figure 1**).

**Table 1.**
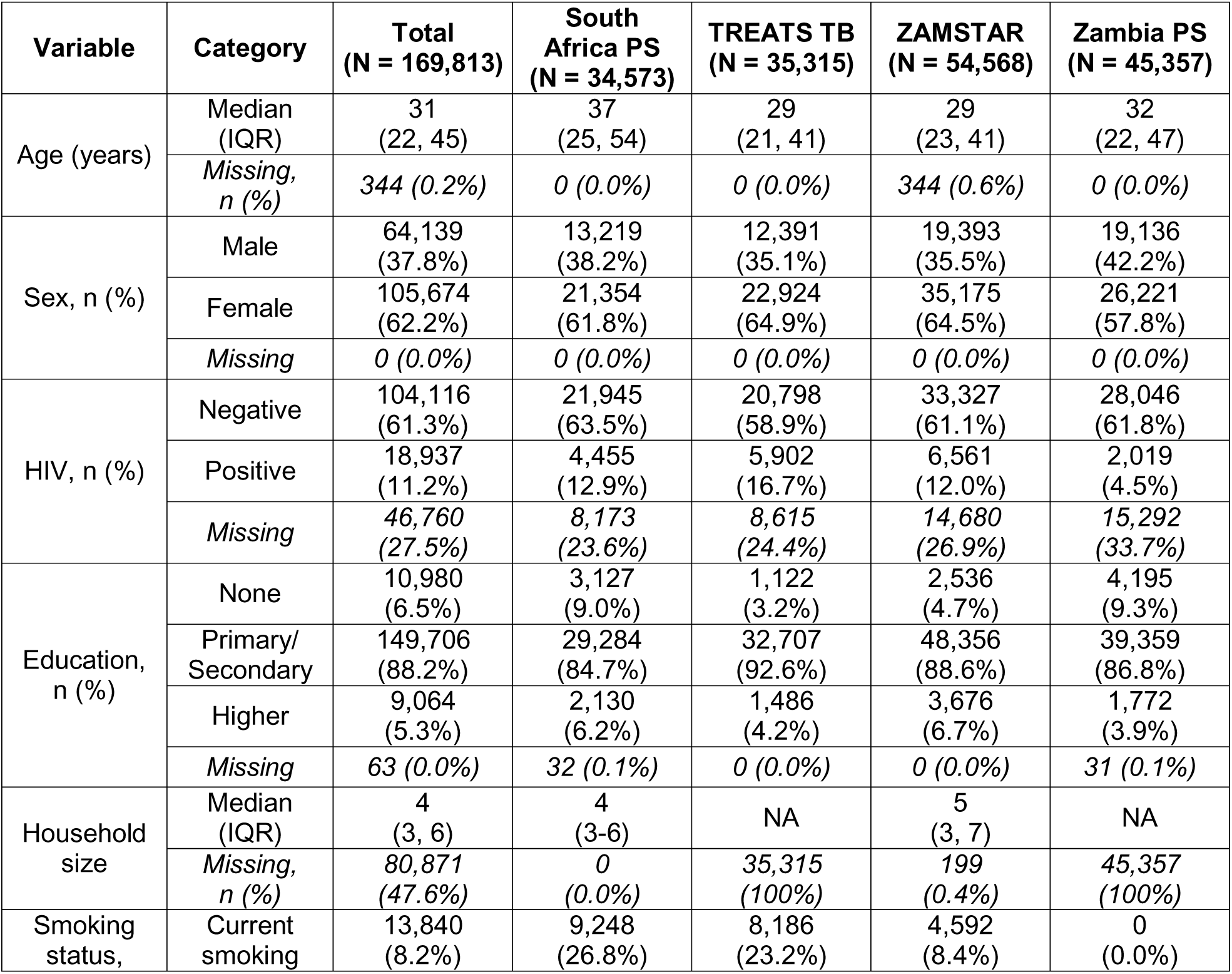
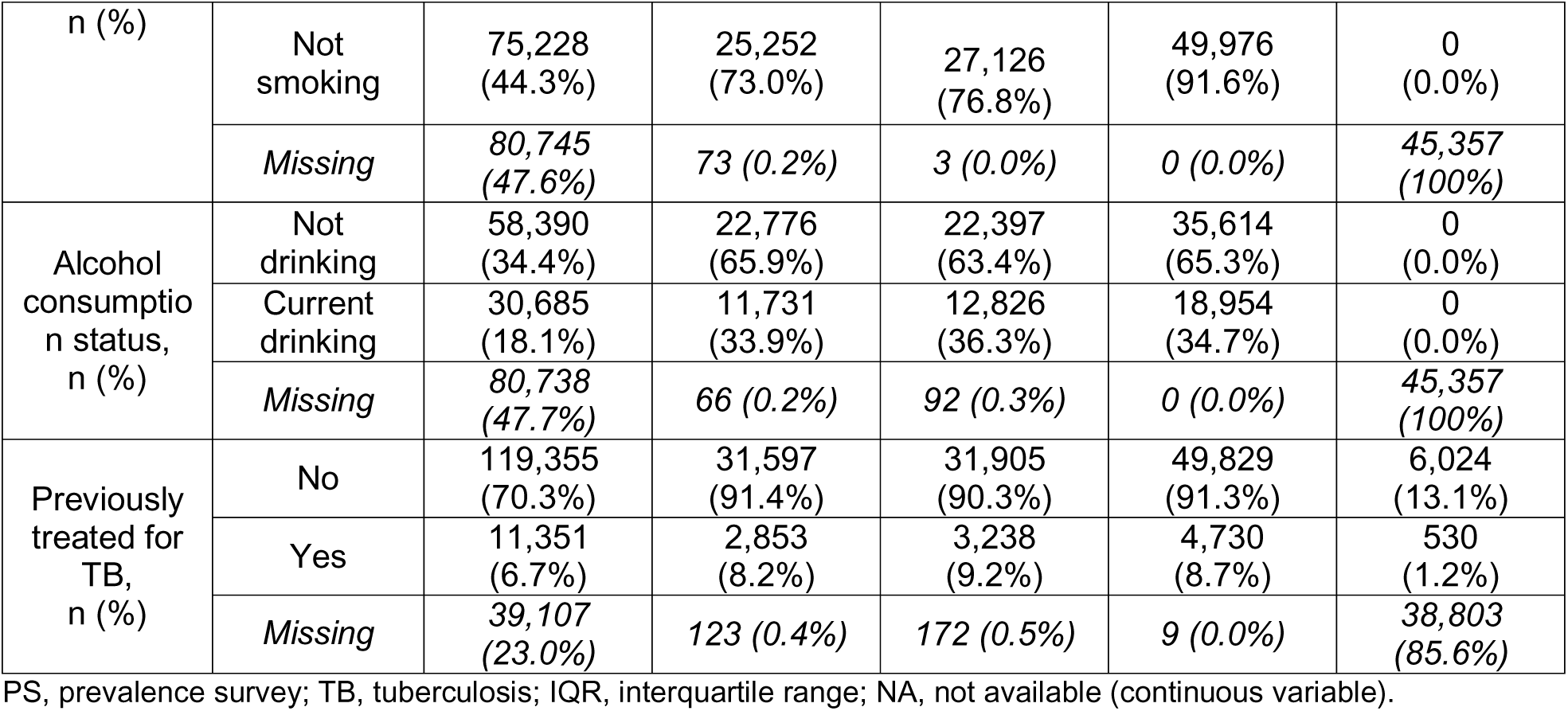
The unimputed participant characteristics by study.

### XGBoost results

Overall discriminative ability reached an AUC of 79.7% (95% CI: 78.7, 80.7) (**Figure 2**). At the Youden threshold (0.45), the model achieved 70.2% sensitivity (95% CI: 64.0, 75.9) and 75.0% specificity (95% CI: 69.8, 79.6). Meeting the WHO TPP 90% sensitivity target required a 0.29 threshold, yielding 46.2% specificity (95% CI: 42.0, 50.4). Conversely, achieving the 60% specificity target required a 0.35 threshold, resulting in 81.5% sensitivity (95% CI: 77.6, 84.9). The model demonstrated strong rule-out capabilities, maintaining a NPV above 95% across all thresholds. However, the PPV remained consistently lower, ranging from 15.0% to 23.2%. Subgroup analyses confirmed consistent discriminatory performance across all clinical strata (**Figure S4**). Calibration assessments indicated acceptable overall accuracy (Brier score=0.16 [95% CI: 0.15, 0.18]) but revealed a systematic overestimation of TB risk (calibration intercept=-2.08 [95% CI: −2.27, −1.88]; calibration slope=1.17 [95% CI: 1.13, 1.22]) requiring potential recalibration depending on programmatic priorities and capacity (**Figure S3**).

**Figure 2.**
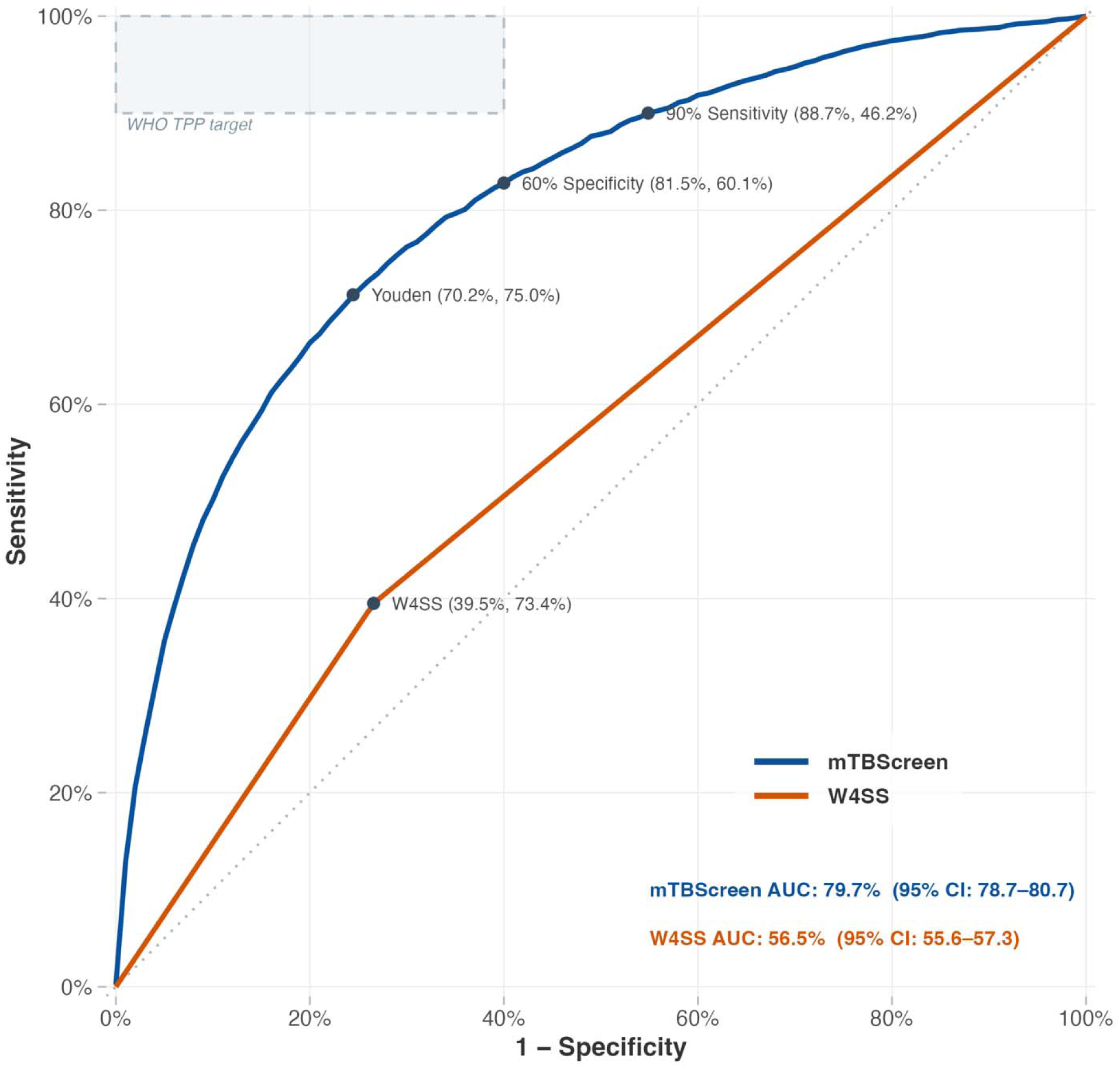
Area under curve (AUC) analysis of XGBoost model. The solid blue line plots the model’s trade-off between sensitivity (Y-axis) and 1-specificity (X-axis) across all possible thresholds. The dashed line represents a non-discriminatory model (AUC = 50%). Grey dots highlight performance at specific decision thresholds: the binary classification for W4SS, the Youden index, and thresholds selected to meet WHO target product profile for a high-sensitivity (90%), moderate-specificity (60%) initial screening tool in a two-step screening approach. WHO, World Health Organization; TPP, Target Product Profile; W4SS. WHO four symptom screen

### Model explainability

SHAP analysis identified age, previous TB treatment, times treated for TB, occupation, and chest pain duration as the top five global predictors (**Figure 3**). High values for these features consistently drove predictions toward the “Possible TB” class (SHAP value > 0). Older age, prior TB history, times treated for TB and unemployment were highly ranked drivers. Key clinical indicators, such as night sweats, cough, and positive HIV status, also exhibited local feature effects increasing the likelihood of a “Possible TB” prediction, albeit with lower global impact.

**Figure 3.**
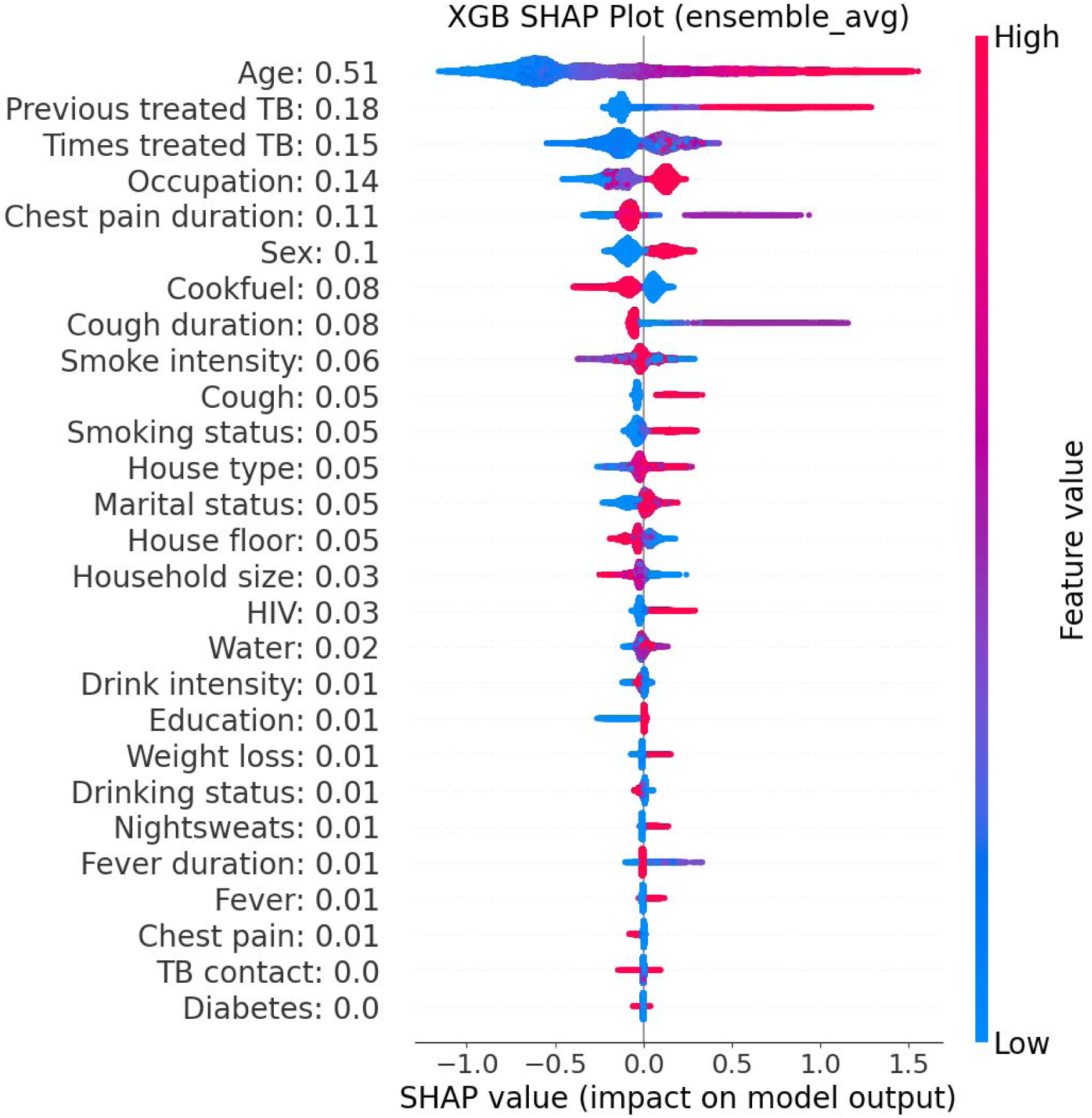
SHapley Additive exPlanations (SHAP) violin plot for model prediction on test data. The plot provides a global summary of each feature’s contribution and effect distribution on the model prediction. The x-axis represents the shapely value magnitude and direction: a value greater than zero indicate that the feature has an impact of driving the model’s prediction of the positive class (Possible TB), whereas values below zero indicate the impact of the model’s prediction of the negative class (Unlikely TB). The colour scheme on the plot is used to map the actual feature value for each observation from the lowest value (blue) to the high value (red).

### XGBoost vs. W4SS

The XGBoost model demonstrated superior diagnostic performance compared to the W4SS across key metrics (**Table 2**), achieving an AUC of 79.7% (95% CI: 78.7, 80.7) versus 57.0% (95% CI: 56.1, 57.9) for the W4SS. At the 0.45 Youden threshold, the ML model significantly outperformed the W4SS, demonstrating higher sensitivity (70.2% [95% CI: 64.0, 75.9] vs. 38.2% [95% CI: 36.6, 40.0]) and nearly double the PPV (23.2% [95% CI: 21.0, 25.4] vs. 14.4% [95% CI: 13.7, 15.2]). Applying the clinically meaningful 60% specificity target (threshold 0.35) yielded a sensitivity more than double that of the W4SS.

**Table 2.** Performance metrics of the XGBoost model and W4SS on the test set.

| Model: Threshold | Metric | Precision, % (95% CI) |
| --- | --- | --- |
| XGBoost:<br>0.45 (Youden) | Sensitivity | 70.2 (64.0, 75.9) |
|  | Specificity | 75.0 (69.8, 79.6) |
|  | PPV | 23.2 (21.0, 25.4) |
|  | NPV | 95.9 (95.3, 96.5) |
| XGBoost:<br>0.29 (90% sensitivity) | Sensitivity | 88.7 (86.4, 90.7) |
|  | Specificity | 46.2 (42.0, 50.4) |
|  | PPV | 15.0 (14.1, 15.9) |
|  | NPV | 97.5 (97.1, 97.8) |
| XGBoost:<br>0.35 (60% specificity) | Sensitivity | 81.5 (77.6, 84.9) |
|  | Specificity | 60.1 (54.8, 65.2) |
|  | PPV | 17.9 (16.5, 19.4) |
|  | NPV | 96.8 (96.4, 97.2) |
| W4SS | Sensitivity | 38.2 (36.6, 40.0) |
|  | Specificity | 75.8 (75.3, 76.3) |
|  | PPV | 14.4 (13.7, 15.2) |
|  | NPV | 92.0 (91.6, 92.3) |

CI, confidence interval; TB, tuberculosis; W4SS, WHO-four symptom screen; PPV, positive predictive value; NPV, negative predictive value
Fixed value based on the assigned threshold.

Decision curve analysis confirmed the clinical utility of the XGBoost model (Figure 4). The model exhibited a positive net benefit across a wide range of threshold probabilities (approximately 0.02 to 0.58), demonstrating superiority over both the “Test All” and “Test None” strategies. Furthermore, the ML model consistently outperformed the W4SS, maintaining a higher net benefit curve across the entire clinically relevant range. Subgroup analyses revealed this clinical utility was particularly pronounced among PLWHIV, previously treated individuals, symptomatic (W4SS+) participants, and males (**Figure S4**).

**Figure 4.**
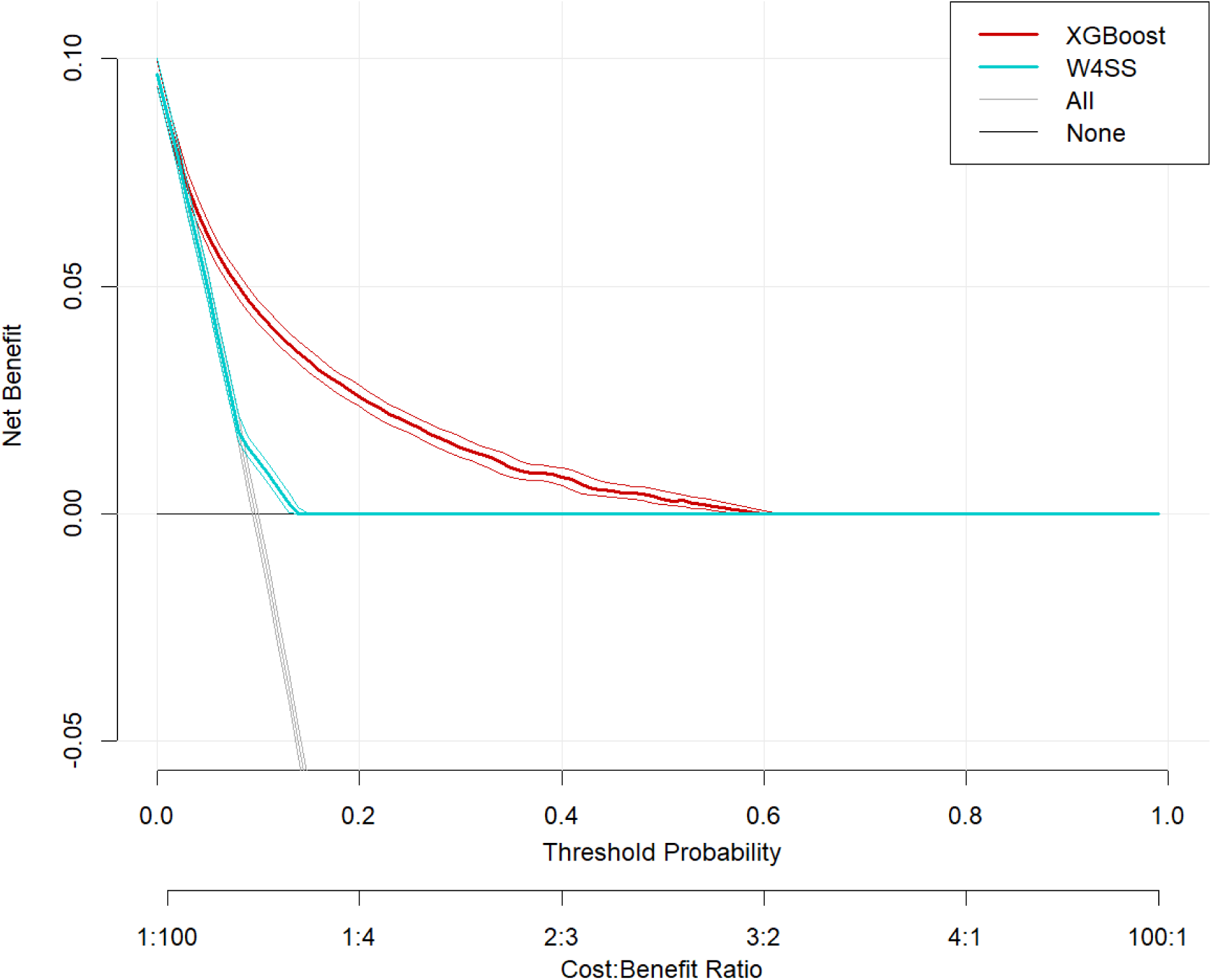
Decision curve analysis (DCA) for the XGBoost model. The plot compares the net benefit (Y-axis) of the ML model (red line, with 95% confidence intervals) against the W4SS algorithm (cyan line). Both are benchmarked against the default strategies of referring all individuals for confirmatory testing (“All”, grey line) and referring none (“None”, black line). The net benefit for each strategy is plotted across a range of threshold probabilities (primary X-axis) and their corresponding Harm:Benefit ratios (secondary X-axis).

## Discussion

This study demonstrates the potential of an ML-based TB screening model. The model was developed using a harmonized dataset derived from four prevalence surveys (two national and two community-based) in South Africa and Zambia, which are representative high-burden settings. When evaluated against a test set, the model achieved good overall discriminative ability (AUC 79.7%). Assessed against WHO TPP criteria, the model did not meet the 90% sensitivity target without a compromise in specificity, but at 60% specificity it achieved 81.5% sensitivity (the WHO target performance is 90% sensitivity and 60% specificity).^12^ Conceivably, as an initial screening test, the ML model could serve as an accessible and low-cost tool to rule out a large proportion of assessed individuals. This would allow TB programs to focus more resource-intensive and less accessible screening tests, such as CXR (possibly with CAD), on a high-probability cohort, thereby increasing the feasibility of large-scale screening programs and optimizing resource allocation.

Decision curve analysis further demonstrated positive clinical utility for the ML-based model. Importantly, this benefit was greatest at lower risk thresholds. This is a clinically important area where decision-making is often difficult, as clinicians must weigh the risk of missing true TB cases against the burden of unnecessary testing. Within this low-threshold range, the model consistently outperformed the W4SS, indicating superior clinical utility for screening and early case detection.

These analyses confirm the poor performance of the W4SS (AUC 57.0%, sensitivity 38.2%) in community settings. By incorporating a wider array of variables (demographics, socio-economic indicators, HIV status, and symptom duration) alongside the four classic symptoms, the ML model increases the discriminative ability. At a threshold achieving the WHO TPP target of 60% specificity, the ML model provides substantial gains in sensitivity (82.3% vs. 38.2%) and modest gains in negative predictive value (96.9% vs. 92.0%) compared to the W4SS. This suggests that leveraging additional, easily collected data within an ML framework creates a more efficient screening tool.

Despite these gains, model accuracy remains inferior to CXR with CAD, which consistently demonstrates superior sensitivity (86.0% to 89.0%) and specificity (59.0% to 80.0%).^30^ However, unlike CXR, which requires significant infrastructure and capital, this model is designed for integration within a custom smartphone application *mTBScreen* (**Figure 5**) intended for decentralized, community-level deployment. Its intended utility is to function as a high-throughput screening tool to rule out low-risk individuals, identifying a smaller, higher-probability cohort to undergo a second screening step with a more accurate tool such as CXR with CAD.

**Figure 5.**
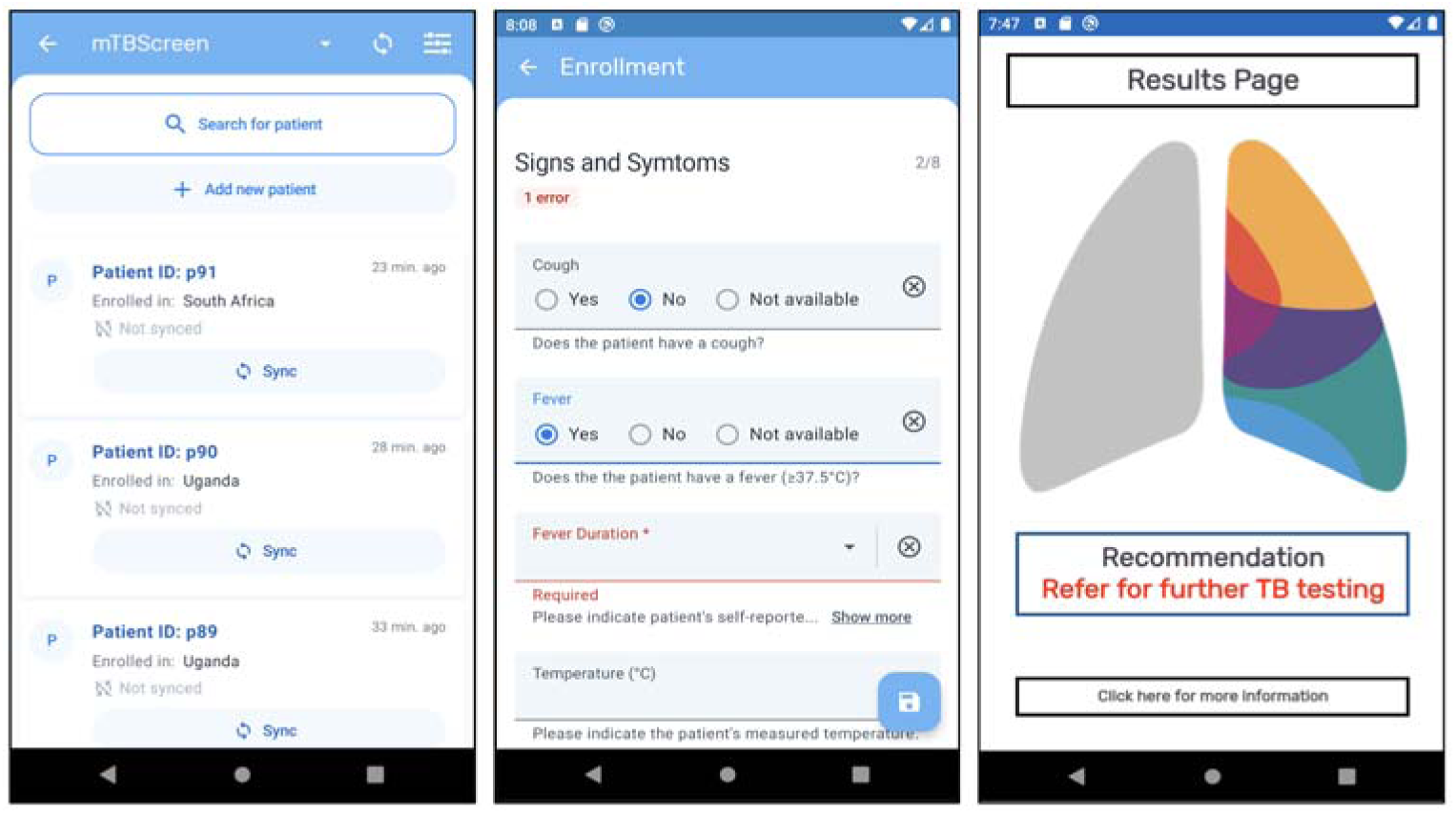
Screenshots of the mTBScreen app. Left: Overview of patient profiles. Middle: Collection form for input patient clinical and demographic variables. Right: Recommendations based on results from TB risk prediction algorithm (operating behind the scenes).

This study has several notable strengths. First, it addresses a timely and relevant topic by applying ML to TB screening, which is an area of importance for improving early detection strategies. In light of the recently approved near point-of-care molecular tests,^31^ an ML app-based screening tool can identify people at high risk of TB for confirmatory testing in peripheral health-care settings. Second, the analysis utilized large, community-based datasets rather than facility-based samples, specifically targeting underdiagnosed populations. Lastly, the model’s performance was directly compared to an established standard, the W4SS, enabling a clear assessment of its added clinical value relative to current practice.

Several limitations should be noted. First, the harmonized dataset was highly imbalanced (9.7% “Possible TB”), which is typical of TB prevalence data. Although inverse proportional weighting managed this during training, the low precision for the positive class (23.5%) indicates the model still produces a high number of false positives, reflecting a common challenge in rare-event prediction. Second, managing missing data represented a methodological challenge. While multiple imputation is widely recommended for handling incomplete data, the XGBoost method lacks standardized procedures for fitting a single unified model. However, this was addressed by utilizing an ensemble approach across the imputed datasets and applying Rubin’s rules with bootstrap resampling to ensure robust confidence intervals.^26^ Third, the outcome definition relied on a composite reference standard that varied across the four source datasets, incorporating different combinations of microbiological and radiological results. This heterogeneity could introduce noise and inconsistency into model predictions. Consequently, benchmarking the model against WHO TPP targets represents a conservative evaluation for sensitivity. Because the composite outcome includes radiographic abnormalities, the model’s sensitivity is penalized when compared to strict TPP thresholds designed solely for microbiologically active disease. Finally, the harmonization process itself posed challenges, as the four datasets were not originally designed to be interoperable. This led to heterogeneity in the definition and availability of input variables and resulted in high missingness for key predictors (e.g., smoking status, household size). For example, the “TB contact” variable had a low global impact on predictions, diverging from established epidemiology where contact is a primary risk factor. This skewed effect likely stems from significant missing data and variable definitions for this variable combined with the intrinsic limitations of using a binary variable to capture complex exposure dynamics.

Future efforts to refine the XGBoost model should prioritize external validation and the integration of high-resolution metadata to enhance clinical utility. Validating the model using independent datasets from diverse geographic regions is essential to ensure its discriminatory performance remains robust and generalizable beyond the current harmonized cohort. Furthermore, predictive power could be improved by incorporating geospatial data to identify community transmission hotspots,^32,33^ thereby accounting for environmental risks often missed by self-reporting alone.

In conclusion, the TB risk prediction XGBoost model represents a promising development in the search for a more accurate and accessible TB screening tool. By leveraging ML on readily available clinical and demographic data, the model outperforms the W4SS, driven largely by its ability to detect high-risk individuals who are asymptomatic and thus missed by standard symptom screening. While this study establishes a strong foundation, future research should focus on prospective validation in diverse geographic regions, evaluating the tool’s cost-effectiveness compared to alternative screening algorithms, identifying novel predictive covariates not currently captured in standard surveys and integrating known other variables, e.g. geolocation. Ultimately, this work demonstrates the potential of ML-based smartphone tools to enhance active case finding, improve screening efficiency, and help identify the undetected millions living with TB.

## List of abbreviations

ACF: Active Case Finding
AUC: Area Under the Curve
CAD: Computer Aided Detection
CAD4TB: Computer Aided Detection for Tuberculosis
CI: Confidence Interval
CXR: Chest X-ray
DCA: Decision Curve Analysis
IQR: Interquartile Range
ML: Machine Learning
PLWHIV: People Living with HIV
SHAP: SHapley Additive exPlanations
TB: Tuberculosis
TPP: Target Product Profile
TREATS: Tuberculosis Reduction through Expanded Anti-retroviral Treatment and Screening
W4SS: World Health Organization Four Symptom Screen
WHO: World Health Organization
XGBoost: eXtreme Gradient Boosting
ZAMSTAR: Zambia, South Africa Tuberculosis and AIDS Reduction

## Contributors

Conceptualization (AK, LMH, SY, CMD), original data acquisition (LK, HA, YH, NK, PK, NK, IL, SM, TM, MoM, AS), data curation (AJZ, KFM, HL, LK, EC, YH, IL, MiM), formal analysis (AJZ, KFM, HL, LK, EC, MO), funding acquisition (LMH, CMD), methodology (AJZ, KFM, HL, LK, EC, MG, KK, LMH, FM, JR, MO, CMD), supervision (CMD), software (CI), visualization (AJZ, KFM, HL), writing-original draft (AJZ, KFM, HL, CMD), writing-review & editing (all authors contributed equally)

## Data Availability

This study is a secondary analysis of existing datasets. The data underlying this article cannot be shared publicly due to ethical and privacy restrictions stipulated by the original data sharing agreements. Researchers wishing to access the raw data must apply directly to the data stewards of the respective source studies.

## Acknowledgements

The authors gratefully acknowledge the data storage service SDS@hd supported by the Ministry of Science, Research and the Arts Baden-Württemberg (MWK) and the German Research Foundation (DFG) through grant INST 35/1503-1 FUGG.

## Data and code availability

This study is a secondary analysis of existing datasets. The data underlying this article cannot be shared publicly due to ethical and privacy restrictions stipulated by the original data sharing agreements. Researchers wishing to access the raw data must apply directly to the data stewards of the respective source studies. The analytical code and model weights developed for this study are currently restricted, as they are actively being integrated into the proprietary mTBScreen smartphone application.

