## Supplementary Information for "Development and validation of a machine learning model for community-based tuberculosis screening among persons aged ≥ 15 years in South Africa and Zambia"

**Table S1.** TRIPOD+AI checklist.

| **Section/Topic** | **Item** | **Checklist item** | **Reported on section** |
| --- | --- | --- | --- |
| **TITLE** | | | |
| *Title* | 1 | Identify the study as developing or evaluating the performance of a multivariable prediction model, the target population, and the outcome to be predicted | Title page. Title identifies the development and validation of an ML model for TB screening among adults >=15 in specific countries. |
| **ABSTRACT** | | | |
| *Abstract* | 2 | See TRIPOD+AI for Abstracts checklist | Abstract. Structured abstract covers development/ validation, population, methods, results, and conclusion. |
| **INTRODUCTION** | | | |
| *Background* | 3a | Explain the healthcare context (including whether diagnostic or prognostic) and rationale for developing or evaluating the prediction model, including references to existing models | Introduction, paragraphs 1 and 2. Explains the need for active case finding and references the limitations of the existing W4SS model. |
|  | 3b | Describe the target population and the intended purpose of the prediction model in the context of the care pathway, including its intended users (e.g., healthcare professionals, patients, public) | Introduction, paragraph 4. Specifies the generation of personalized TB risk scores for individuals aged >=15 years in community settings. |
|  | 3c | Describe any known health inequalities between sociodemographic groups | Introduction, paragraph 1. Touches on structural barriers preventing timely care for underdiagnosed populations. |
| *Objectives* | 4 | Specify the study objectives, including whether the study describes the development or validation of a prediction model (or both) | Introduction, paragraph 4. Specifies the aim to develop and validate the ML prediction model. |
| **METHODS** | | | |
| *Data* | 5a | Describe the sources of data separately for the development and evaluation datasets, the rationale for using these data, and representativeness of the data | Methods, Datasets and population. Describes the four national and community-based TB prevalence surveys analyzed |
|  | 5b | Specify the dates of the collected participant data, including start and end of participant accrual; and, if applicable, end of follow-up | Methods, Datasets and population & Table S2. Lists the specific collection years for all four surveys (2010 to 2021). |
| *Participants* | 6a | Specify key elements of the study setting (e.g., primary care, secondary care, general population) including the number and location of centres | Methods, Datasets and population & Table S2. Details the community-based survey settings across specific clusters and provinces. |
|  | 6b | Describe the eligibility criteria for study participants | Methods, Datasets and population. Defines inclusion criteria (adults >=15) and exclusion criteria (on TB treatment or missing test results). |
|  | 6c | Give details of any treatments received, and how they were handled during model development or evaluation, if relevant | Methods, Datasets and population. Explicitly notes that participants currently receiving TB treatment were excluded. |
| *Data preparation* | 7 | Describe any data pre-processing and quality checking, including whether this was similar across relevant sociodemographic groups | Methods, Input variables for model development. Explains that variables were retained and harmonized if present in at least two sources with <20% missing values. |
| *Outcome* | 8a | Clearly define the outcome that is being predicted and the time horizon, including how and when assessed, the rationale for choosing this outcome, and whether the method of outcome assessment is consistent across sociodemographic groups | Methods, Outcome definition for model training. Defines the composite reference standard used to classify "Possible TB" versus "Unlikely TB". |
|  | 8b | If outcome assessment requires subjective interpretation, describe the qualifications and demographic characteristics of the outcome assessors | N/A |
|  | 8c | Report any actions to blind assessment of the outcome to be predicted | N/A |
| *Predictors* | 9a | Describe the choice of initial predictors (e.g., literature, previous models, all available predictors) and any pre-selection of predictors before model building | Methods, Input variables for model development. States the 27 variables were chosen based on known TB associations and ease of measurement. |
|  | 9b | Clearly define all predictors, including how and when they were measured (and any actions to blind assessment of predictors for the outcome and other predictors) | Methods, Input variables for model development & Tables S4/S5. Summarizes predictor harmonization and missingness thresholds. |
|  | 9c | If predictor measurement requires subjective interpretation, describe the qualifications and demographic characteristics of the predictor assessors | Methods, Input variables for model development. Predictors are largely self-reported survey data. |
| *Sample size* | 10 | Explain how the study size was arrived at (separately for development and evaluation), and justify that the study size was sufficient to answer the research question. Include details of any sample size calculation | Methods, Data partitioning, sample size, and missing data imputation, Notes sample size was fixed by available data and confirmed adequate via Riley criteria/pmsampsize. |
| *Missing data* | 11 | Describe how missing data were handled. Provide reasons for omitting any data | Methods, Data partitioning, sample size, and missing data imputation. Handled using multiple imputation by chained equations (MICE). |
| *Analytical methods* | 12a | Describe how the data were used (e.g., for development and evaluation of model performance) in the analysis, including whether the data were partitioned, considering any sample size requirements | Methods, Data partitioning, sample size, and missing data imputation. Describes the 80/20 train/test split, stratified by study and outcome. |
|  | 12b | Depending on the type of model, describe how predictors were handled in the analyses (functional form, rescaling, transformation, or any standardisation). | Methods, Data partitioning, sample size, and missing data imputation. Imputed datasets generated via mice package and evaluated on test sets. |
|  | 12c | Specify the type of model, rationale, all model-building steps, including any hyperparameter tuning, and method for internal validation | Methods, Model training and evaluation. Details the XGBoost ensemble model built in Python and Bayesian search cross-validation for hyperparameters.Table S7 provides hyperparameter tuning info. |
|  | 12d | Describe if and how any heterogeneity in estimates of model parameter values and model performance was handled and quantified across clusters (e.g., hospitals, countries). See TRIPOD-Cluster for additional considerations^3^ | Results, XGBoost results & Figure S4. Subgroup analyses performed to confirm consistent performance across strata. |
|  | 12e | Specify all measures and plots used (and their rationale) to evaluate model performance (e.g., discrimination, calibration, clinical utility) and, if relevant, to compare multiple models | Methods, Model training and evaluation. Lists AUC, sensitivity, specificity, predictive values, Brier score, and calibration metrics. |
|  | 12f | Describe any model updating (e.g., recalibration) arising from the model evaluation, either overall or for particular sociodemographic groups or settings | Results, XGBoost results. Notes a systematic overestimation requiring potential future recalibration. |
|  | 12g | For model evaluation, describe how the model predictions were calculated (e.g., formula, code, object, application programming interface) | Note S1 and Note S2 provide equations for outcome metrics. Discussion & Figure 5. Predictions are integrated into a custom smartphone application (mTBScreen). |
| *Class imbalance* | 13 | If class imbalance methods were used, state why and how this was done, and any subsequent methods to recalibrate the model or the model predictions | Methods, Model training and evaluation. Addressed using inverse proportional weighting during training. |
| *Fairness* | 14 | Describe any approaches that were used to address model fairness and their rationale | Methods, Decision curve analysis for clinical utility. Evaluated clinical utility via subgroup analyses for sex, HIV status, etc. |
| *Model output* | 15 | Specify the output of the prediction model (e.g., probabilities, classification). Provide details and rationale for any classification and how the thresholds were identified | Methods, Model training and evaluation. Assessed across three thresholds (Youden index, 90% sensitivity, 60% specificity) to classify "Possible TB". |
| *Training versus*  *evaluation* | 16 | Identify any differences between the development and evaluation data in healthcare setting, eligibility criteria, outcome, and predictors | Methods, Data partitioning, sample size, and missing data imputation & Table S5. Data split was stratified to maintain comparable distributions between sets. |
| **OPEN SCIENCE** | | | |
| *Ethical approval* | 17 | Name the institutional research board or ethics committee that approved the study and describe the participant-informed consent or the ethics committee waiver of informed consent | Methods, Ethics. Approved by the Ethics Committee of the Medical Faculty of Heidelberg University. |
| Funding | 18a | Give the source of funding and the role of the funders for the present study | Funding section. |
| Conflicts of interest | 18b | Declare any conflicts of interest and financial disclosures for all authors | Declarations of interest section. |
| Protocol | 18c | Indicate where the study protocol can be accessed or state that a protocol was not prepared | Abstract, No study registration. |
| Registration | 18d | Provide registration information for the study, including register name and registration number, or state that the study was not registered | This study was not registered. It is a retrospective analysis of data. |
| Data sharing | 18e | Provide details of the availability of the study data | Data and code availability section. |
| Code sharing | 18f | Provide details of the availability of the analytical code | Data and code availability section. |
| **PATIENT & PUBLIC INVOLVEMENT** | | | |
| Patient & Public Involvement | 19 | Provide details of any patient and public involvement during the design, conduct, reporting, interpretation, or dissemination of the study or state no involvement. | N/A |
| **RESULTS** | | | |
| Participants | 20a | Describe the flow of participants through the study, including the number of participants with and without the outcome and, if applicable, a summary of the follow-up time. A diagram may be helpful. | Results, Participant characteristics & Figure 1. Flow diagram detailing inclusion/exclusion steps and final counts. |
|  | 20b | Report the characteristics overall and, where applicable, for each data source or setting, including the key dates, key predictors (including demographics), treatments received, sample size, number of outcome events, follow-up time, and amount of missing data. A table may be helpful. Report any  differences across key demographic groups. | Results, Participant characteristics & Tables 1/S3. Summarizes participant characteristics, demographics, and missing data. |
|  | 20c | For model evaluation, show a comparison with the development data of the distribution of important predictors (demographics, predictors, and outcome). | Supplementary Table S6. Compares the distribution of predictors across training and test data splits. |
| Model development | 21 | Specify the number of participants and outcome events in each analysis (e.g., for model development, hyperparameter tuning, model evaluation) | Methods, Data partitioning, sample size, and missing data imputation & Results, Participant characteristics. Details sample sizes for training/eval and the final number of Possible TB events. Table S2 shows distribution by data source. |
| Model specification | 22 | Provide details of the full prediction model (e.g., formula, code, object, application programming interface) to allow predictions in new individuals and to enable third-party evaluation and implementation, including any restrictions to access or re-use (e.g., freely available, proprietary)^5^ | Supplementary Table S7 & Discussion (Figure 5). Lists hyperparameters and shows the app interface intended for predictions. |
| Model performance | 23a | Report model performance estimates with confidence intervals, including for any key subgroups (e.g., sociodemographic). Consider plots to aid presentation. | Results, XGBoost results & Table 2 & Figure 2. Reports AUC, sensitivity, specificity, PPV, NPV with 95% CIs. |
|  | 23b | If examined, report results of any heterogeneity in model performance across clusters. See TRIPOD Cluster for additional details3. | Results, XGBoost results & Supplementary Figure S4. Confirms performance across clinical strata subgroups. |
| Model updating | 24 | Report the results from any model updating, including the updated model and subsequent performance | Results, XGBoost results. Model was not updated, but recalibration was flagged as a potential future need. |
| **DISCUSSION** | | | |
| Interpretation | 25 | Give an overall interpretation of the main results, including issues of fairness in the context of the objectives and previous studies | Discussion, paragraphs 1 to 3. Evaluates the ML model's potential and clinical utility compared to the W4SS standard. |
| Limitations | 26 | Discuss any limitations of the study (such as a non-representative sample, sample size, overfitting, missing data) and their effects on any biases, statistical uncertainty, and generalizability | Discussion, paragraphs 6 to 9. Acknowledges issues like class imbalance, missing data handling, and composite reference standards. |
| Usability of the model in the context of current care | 27a | Describe how poor quality or unavailable input data (e.g., predictor values) should be assessed and handled when implementing the prediction model | Discussion, paragraph 9. Explains that data heterogeneity resulted in high missingness for predictors like TB contact, impacting model logic. |
|  | 27b | Specify whether users will be required to interact in the handling of the input data or use of the model, and what level of expertise is required of users | Discussion, paragraph 5 & Figure 5. Model is built into a smartphone app designed for decentralized deployment. |
|  | 27c | Discuss any next steps for future research, with a specific view to applicability and generalizability of the model | Discussion, paragraphs 10 to 11. Suggests external validation and the addition of geospatial data to account for environmental risks. |

TRIPOD=Transparent Reporting of a multivariable prediction model for Individual Prognosis Or Diagnosis; AI=artificial intelligence

**Table S2.** Data sources for TB risk prediction model training and validation.

| **Data source** | **Country** | **Coverages** | **Collection year** | **Eligible (N)^*^** | **Possible TB^†^ (%)** | **Unlikely TB^†^ (%)** |
| --- | --- | --- | --- | --- | --- | --- |
| South Africa PS (WHO) | South Africa | 110 clusters across all 9 provinces | 2017-2019 | 34,573 | 5,770 (16.7) | 28,803 (83.3) |
| Zambia PS (WHO) | Zambia | 66 clusters across all 10 provinces | 2013-2014 | 45,357 | 4,028 (8.9) | 41,329 (91.1) |
| TREATS TB PS (Zambart) | South Africa & Zambia | 21 urban and peri-urban communities | 2019-2021 | 35,315 | 4,568 (12.9) | 30,747 (87.1) |
| ZAMSTAR clinical trial (Zambart) | South Africa & Zambia | 24 urban and rural communities | 2010 | 54,568 | 2,047 (3.8) | 52,521 (96.2) |
| **Total** | | | | **169,813** | **16,413 (9.7)** | **153,400 (90.3)** |

WHO, World Health Organization; PS, prevalence survey; TREATS TB, Tuberculosis Reduction through Expanded Antiretroviral Treatment and Screening for Active Tuberculosis; ZAMSTAR, Zambia, South Africa Tuberculosis and AIDS Reduction; N; number; TB, tuberculosis

^*^Eligibility based on age (adults ≥15 years), not currently under active TB treatment, and availability of TB X-ray and/or diagnostic results.

^†^Based on composite reference defined within the mTBScreen study.

**Table S3.** Outcome definitions by each study.

|  | **Possible TB** | | **Unlikely TB** | |
| --- | --- | --- | --- | --- |
| **Study** | **Definition** | **N (%)** | **Definition** | **N (%)** |
| South Africa PS | Either:   - Sputum culture positive OR - Sputum Xpert positive OR - Abnormal CXR | 5,770 (16.7) | Not TB Screen Positive and at least one of:   - Sputum culture negative OR - Normal CXR OR - Sputum Xpert negative | 28,803 (83.3) |
| Zambia PS | Either:   - Sputum culture positive^*^ OR - Sputum Xpert positive OR - Smear microscopy positive OR - Abnormal CXR | 4,028 (8.9) | Not TB Screen Positive and at least one of:   - Sputum culture negative OR - Sputum Xpert negative OR - Smear microscopy negative OR - Normal CXR | 41,329 (91.1) |
| TREATS TB | Either:   - Sputum Xpert positive OR - Smear microscopy positive OR - Abnormal CXR - CAD score ≥50^†^ | 4,568 (12.9) | Not TB Screen Positive and at least one of:   - Sputum Xpert negative OR - Smear microscopy negative OR - Normal CXR OR - CAD score <50^†^ | 30,747 (87.1) |
| ZAMSTAR | Either:   - Sputum culture positive^*^ OR - Sputum microscopy positive | 2,047 (3.8) | Not TB Screen Positive and at least one of:   - Sputum culture negative OR - Smear microscopy negative OR | 52,521 (96.2) |
| **Total** |  | 16413 (9.7) |  | 153,400 (90.3) |

^*^Includes positive for both TB and nontuberculous mycobacteria.

^†^The cut-off reported in the TREATS TB study.

PS, prevalence survey; TREATS TB, Tuberculosis Reduction through Expanded Antiretroviral Treatment and Screening for Active Tuberculosis; ZAMSTAR, Zambia, South Africa Tuberculosis and AIDS Reduction

Both NTM and MTB were considered as “Possible TB” in ZAMSTAR due to the overlapping clinical presentation of TB and NTM.

**Table S4.** Summary of unimputed variables used as inputs in the XGBoost model by data source.

| **Variable** | **Category** | **South Africa PS**  **(N = 34,573)** | **TREATS TB (N = 35,315)** | **ZAMSTAR (N = 54,568)** | **Zambia PS (N = 45,357)** | **Total**  **(N = 169,813)** |
| --- | --- | --- | --- | --- | --- | --- |
| Age (years) | Median (IQR) | 37  (25-54) | 29  (21, 41) | 29  (23, 41) | 32  (22, 47) | 31  (22, 45) |
|  | *Missing* | *0 (0.0)* | *0 (0.0)* | *344 (0.6)* | *0 (0.0)* | *344 (0.2)* |
| Sex | Male | 13,219 (38.2) | 12,391 (35.1) | 19,393 (35.5) | 19,136 (42.2) | 64,139 (37.8) |
|  | Female | 21,354 (61.8) | 22,924 (64.9) | 35,175 (64.5) | 26,221 (57.8) | 105,674 (62.2) |
|  | *Missing* | *0 (0.0)* | *0 (0.0)* | *0 (0.0)* | *0 (0.0)* | *0 (0.0)* |
| Marital status | Single | 0 (0.0) | 19,957 (56.5) | 26,779 (49.1) | 12,812 (28.2) | 59,548 (35.1) |
|  | Married | 0 (0.0) | 11,491 (32.5) | 22,214 (40.7) | 25,664 (56.6) | 59,369 (35.0) |
|  | Separated/  divorced/widowed | 0 (0.0) | 3,867 (11.0) | 5,575 (10.2) | 6,881 (15.2) | 16,323 (9.6) |
|  | *Missing* | *34,573*  *(100)* | *0 (0.0)* | *0 (0.0)* | *0 (0.0)* | *34,573*  *(20.4)* |
| Education | None | 3,127  (9.0) | 1,122  (3.2) | 2,536  (4.7) | 4,195  (9.3) | 10,980  (6.5) |
|  | Primary/ Secondary | 29,284 (84.7) | 32,707 (92.6) | 48,356 (88.6) | 39,359 (86.8) | 149,706 (88.2) |
|  | Higher | 2,130  (6.2) | 1,486  (4.2) | 3,676  (6.7) | 1,772  (3.9) | 9,064  (5.3) |
|  | *Missing* | *32 (0.1)* | *0 (0.0)* | *0 (0.0)* | *31 (0.1)* | *63 (0.0)* |
| Occupation | Employed full time | 5,521  (16.0) | 3,497  (9.9) | 19,176  (35.1) | 4,038  (8.9) | 32,232  (19.0) |
|  | Part-time/informal | 1,907  (5.5) | 6,671  (18.9) | 9,028  (16.5) | 20,485(45.2) | 38,091(22.4) |
|  | Unemployed | 21,974  (63.6) | 19,967  (56.5) | 18,374  (33.7) | 13,993  (30.9) | 74308  (43.8) |
|  | Student | 4,950  (14.3) | 4,638  (13.1) | 7,990  (14.6) | 6377  (14.1) | 23,955  (14.1) |
|  | *Missing* | *221 (0.6)* | *542 (1.5)* | *0 (0.0)* | *464 (1.0)* | *1,227 (0.7)* |
| Smoking status | Current smoking | 9,248  (26.8) | 8,186  (23.2%) | 4,592  (8.4) | 0  (0.0) | 13,840  (8.2) |
|  | Not smoking | 25,252 (73.0) | 27,126  (76.8%) | 49,976 (91.6) | 0  (0.0) | 75,228 (44.3) |
|  | *Missing* | *73 (0.2)* | *3 (0.0%)* | *0 (0.0)* | *45,357 (100)* | *80,745 (47.6)* |
| Smoke intensity | Not smoking | 25,252 (73.0) | 27,126 (76.8%) | 49,976 (91.6) | 0  (0.0) | 75,228 (44.3) |
|  | Low | 492(1.4) | 1,547 (4.4%) | 443  (0.8) | 0  (0.0) | 935  (0.6) |
|  | High | 8,676 (25.1) | 6,639  (18.8%) | 3,206  (5.9) | 0  (0.0) | 11,882 (7.0) |
|  | *Missing* | *153 (0.4)* | *3 (0.0%)* | *943 (1.7)* | *45,357 (100)* | *81,768 (48.2)* |
| Drinking status | Not drinking | 22,776 (65.9) | 22,397 (63.4%) | 35,614 (65.3) | 0  (0.0) | 58,390 (34.4) |
|  | Currently drinking | 11,731 (33.9) | 12,826  (36.3%) | 18,954 (34.7) | 0  (0.0) | 30,685 (18.1) |
|  | *Missing* | *66 (0.2)* | *92 (0.3%)* | *0 (0.0)* | *45,357 (100)* | *80,738 (47.7)* |
| Drink intensity | Not drinking | 22,776 (65.9) | 22,397  (63.4%) | 35,614 (65.3) | 0  (0.0) | 58,390 (34.4) |
|  | Occasional | 10,922 (31.6) | 12,002  (34.0%) | 17,060 (31.3) | 0  (0.0) | 27,982 (16.5) |
|  | Heavy | 802  (2.3) | 824  (2.3) | 1,894  (3.5) | 0  (0.0) | 2,696  (1.6) |
|  | *Missing* | *73 (0.2)* | *92 (0.3%)* | *0 (0.0)* | *45,357 (100)* | *80,745 (47.6)* |
| Household size | Median (IQR)  *Missing* | 4 (3-6) | NA | 5 (3-7) | NA | 4 (3-6) |
|  |  | *0 (0.0)* | *35,315 (100%)* | *199 (0.4%)* | *45,357*  *(100%)* | *80,871*  *(47.6%)* |
| House type | Single unit/brick | 0  (0.0) | 17,192 (48.7) | 29,236 (53.6) | 21,958 (48.4) | 68,386 (40.3) |
|  | Cluster/multi-unit/mudadadas | 0  (0.0) | 8,887 (25.2) | 8,176 (15.0) | 1,395  (3.1) | 18,458 (10.9) |
|  | Flat in block of flats | 0  (0.0) | 3,909 (11.1) | 5,028  (9.2) | 1,503  (3.3) | 10,440 (6.2) |
|  | Traditional hut/structure | 0  (0.0) | 152  (0.4) | 8,499 (15.6) | 14,558 (32.1) | 23,209 (13.7) |
|  | Servant quarters or informal dwelling | 0  (0.0) | 5,129 (14.5) | 1,483  (2.7) | 151  (0.3) | 6,763  (4.0) |
|  | *Missing* | *34,573 (100)* | *46 (0.1)* | *2,146 (3.9)* | *5,792 (12.8)* | *42,557 (25.1)* |
| House floor | High quality | 0 (0.0) | 10,673 (30.2) | 16,323 (29.9) | 493 (1.1) | 27,489 (16.2) |
|  | Medium quality | 0 (0.0) | 22,907 (64.9) | 28,370 (52.0) | 19,702 (43.4) | 70,979 (41.8) |
|  | Low quality | 0 (0.0) | 1,659 (4.7) | 7,547 (13.8) | 18,584 (41.0) | 27,790 (16.4) |
|  | *Missing* | *34,573 (100)* | *76 (0.2)* | *2,328 (4.3)* | *6,578 (14.5)* | *43,555 (25.6)* |
| Water source | Direct household access | 17,027 (49.2) | 11,658 (33.0) | 18,198 (33.4) | 1,584 (3.5) | 48,467 (28.5) |
|  | On premise | 8,979 (26.0) | 12,586 (35.6) | 14,562 (26.7) | 2,846 (6.3) | 38,973 (23.0) |
|  | External | 7,789 (22.5) | 11,045 (31.3) | 21,519 (39.4) | 35,051 (77.3) | 75,404 (44.4) |
|  | *Missing* | *778 (2.2)* | *26 (0.1)* | *289 (0.5)* | *5,876 (13.0)* | *6,969 (4.1)* |
| Cook fuel | Clean | 30,864 (89.3) | 16,964 (48.0) | 33,518 (61.4) | 4,728 (10.4) | 86,074 (50.7) |
|  | Unclean | 3,064 (8.9) | 18,319 (51.9) | 21,022 (38.5) | 34,832 (76.8) | 77,235 (45.5) |
|  | *Missing* | *647 (1.9)* | *32 (0.1)* | *28 (0.1)* | *5,797 (12.8)* | *6,504 (3.8)* |
| Cough | No | 31,668 (91.6) | 31,742 (89.9) | 48,037 (88.0) | 34,011 (75.0) | 145,458 (85.7) |
|  | Yes | 2,905 (8.4) | 3,573 (10.1) | 6,531 (12.0) | 11,346 (25.0) | 24,355 (14.3) |
|  | *Missing* | *0 (0.0)* | *0 (0.0)* | *0 (0.0)* | *0 (0.0)* | *0 (0.0)* |
| Cough duration | No symptom | 31,668 (91.6) | 31,742 (89.9) | 48,037 (88.0) | 34,011 (75.0) | 145,458 (85.7) |
|  | <2 weeks | 1,364 (4.0) | 2,667 (7.6) | 4,040 (7.4) | 8,468 (18.7) | 16,539 (9.7) |
|  | >=2 weeks | 1,471 (4.2) | 906 (2.6) | 2,339 (4.3) | 2,878 (6.4) | 7,594 (4.5) |
|  | *Missing* | *70 (0.2)* | *0 (0.0)* | *152 (0.3)* | *0 (0.0)* | *222 (0.1)* |
| Fever | No | 33,141 (95.9) | 34,029 (96.4) | 48,210 (88.4) | 38,059 (84.0) | 153,439 (90.4) |
|  | Yes | 1,432 (4.1) | 1,286 (3.6) | 6,358 (11.6) | 7,298 (16.1) | 16,374 (9.6) |
|  | *Missing* | *0 (0.0)* | *0 (0.0)* | *0 (0.0)* | *0 (0.0)* | *0 (0.0)* |
| Fever duration | No symptom | 33,141 (95.9) | 34,029 (96.4) | 0  (0.0) | 38,059 (83.9) | 105,229 (62.1) |
|  | <2 weeks | 749 (2.2) | 1,021 (2.9) | 0 (0.0) | 5,781 (12.8) | 7,551 (4.5) |
|  | >=2 weeks | 631 (1.8) | 265 (0.8) | 0 (0.0) | 1,517 (3.3) | 2,413 (1.4) |
|  | *Missing* | *52 (0.2)* | *0 (0.0)* | *54,568 (100)* | *0 (0.0)* | *54,620 (32.2)* |
| Night sweats | No | 32,515 (94.1) | 33,858 (95.9) | 48,787 (89.4) | 4,199 (9.3) | 119,359 (70.3) |
|  | Yes | 2,058 (6.0) | 1,457 (4.1) | 5,781 (10.6) | 2,372 (5.2) | 11,668 (6.9) |
|  | *Missing* | *0 (0.0)* | *0 (0.0)* | *0 (0.0)* | *38,786 (85.5)* | *38,786 (22.8)* |
| Weight loss | No | 32,998 (95.4) | 33,769 (95.6) | 47,314 (86.7) | 4,324 (9.5) | 118,405 (69.7) |
|  | Yes | 1,575 (4.6) | 1,546 (4.4) | 7,254 (13.3) | 2,247 (5.0) | 12,622 (7.4) |
|  | *Missing* | *0 (0.0)* | *0 (0.0)* | *0 (0.0)* | *38,786 (85.5)* | *38,786 (22.8)* |
| Chest pain | No | 0 (0.0) | 33,004 (93.5) | 49,576 (90.8) | 31,615 (69.7) | 114,195 (67.2) |
|  | Yes | 0 (0.0) | 2,311 (6.5) | 4,992 (9.2) | 13,742 (30.3) | 21,045 (12.4) |
|  | *Missing* | *34,573 (100)* | *0 (0.0)* | *0 (0.0)* | *0 (0.0)* | *34,573 (20.4)* |
| Chest pain duration | No symptom | 0 (0.0) | 33,004 (93.5) | 0 (0.0) | 31,615 (69.7) | 64,619 (38.1) |
|  | <2 weeks | 0 (0.0) | 1,213 (3.4) | 0 (0.0) | 9,935 (21.9) | 11,148 (6.6) |
|  | >=2 weeks | 0 (0.0) | 1,098 (3.1) | 0 (0.0) | 3,807 (8.4) | 4,905 (2.9) |
|  | *Missing* | *34,573 (100)* | *0 (0.0)* | *54,568 (100)* | *0 (0.0)* | *89,141 (52.5)* |
| TB contact | No | 0 (0.0) | 33,563 (95.0) | 52,854 (96.9) | 6,331 (14.0) | 92,748 (54.6) |
|  | Yes | 0 (0.0) | 1,297 (3.7) | 1,714 (3.1) | 577 (1.3) | 3,588 (2.1) |
|  | *Missing* | *34,573 (100)* | *455 (1.3)* | *0 (0.0)* | *38,449 (84.8)* | *73,477 (43.3)* |
| Previously treated for TB | No | 31,597(91.4) | 31,905 (90.3) | 49,829 (91.3) | 6,024  (13.1) | 119,355 (70.3) |
|  | Yes | 2,853  (8.2) | 3,238  (9.2) | 4,730  (8.7) | 530  (1.2) | 11,351  (6.7) |
|  | *Missing* | *123 (0.4)* | *172 (0.5)* | *9 (0.0)* | *38,803 (85.6)* | *39,107 (23.0)* |
| Times treated for TB | Never | 31,597 (91.4) | 0 (0.0) | 49, 829 (91.3) | 0 (0.0) | 81,426 (48.0) |
|  | Once | 2,583 (7.5) | 0 (0.0) | 3,866 (7.1) | 0 (0.0) | 6,449 (3.8) |
|  | Twice+ | 250 (0.7) | 0 (0.0) | 854 (1.6) | 0 (0.0) | 1,104 (0.6) |
|  | *Missing* | *143 (0.4)* | *35,315 (100)* | *19 (0.0)* | *45,357 (100)* | *80,834 (47.6)* |
| HIV | Negative | 21,945 (63.5) | 20,798 (58.9) | 33,327 (61.1) | 28,046 (61.8) | 104,116 (61.3) |
|  | Positive | 4,455  (12.9) | 5,902  (16.7) | 6,561  (12.0) | 2,019  (4.5) | 18,937 (11.2) |
|  | *Missing* | *8,173*  *(23.6)* | *8,615*  *(24.4)* | *14,680 (26.9)* | *15,292 (33.7)* | *46,760 (27.5)* |
| Diabetes | No | 32,307 (93.4) | 0 (0.0) | 52,080 (95.4) | 0 (0.0) | 84,387 (49.7) |
|  | Yes | 1,7371,740 (5.0) | 0 (0.0) | 2,488 (4.6) | 0 (0.0) | 4,225 (2.5) |
|  | *Missing* | *529 (1.5)* | *35,315 (100)* | *0 (0.0)* | *45,357 (100)* | *81,201 (47.8)* |

PS, prevalence survey; TB, tuberculosis; IQR, interquartile range; NA, not available

**Table S5.** Variable harmonization and recoding.

| **Variable** | **Original variable definition / coding** | **Harmonized definition / coding** |
| --- | --- | --- |
| Smoke intensity | Not smoking | Not smoking |
|  | Less than daily | Low |
|  | Daily | High |
| Drink intensity | Not drinking | Not drinking |
|  | <4 times per week | Occasional |
|  | >=4 times per week | Heavy |
| House type | House/brick structure on own stand (single unit) | Single unit/brick |
|  | Cluster / multi-unit | Cluster/multi-unit/mudadadas |
|  | Flat in block of flats | Flat in block of flats |
|  | Traditional welling/hut/structure made from traditional material | Traditional hut/structure |
|  | Servant quarter, Caravan/Tent, worker’s hostel | Servant quarters or informal dwelling |
| House floor | Wood, linoleum, or tile | High quality |
|  | Cement | Medium quality |
|  | Dirt | Low quality |
| Water source | Piped water (tap) in dwelling | Direct household access |
|  | Piped water (tap) in site/yard | On premise |
|  | Water carrier/tanker, rain-water tank, borehole / well / spring, dam/river/stream, public / communal tap | External |
| Cook fuel | No cooking, electric, or gas | Clean |
|  | Paraffin, charcoal, animal dung, or wood | Unclean |
| TB contact | Not self reported TB contact or  Not living in a household with conformed TB positive during the survey | No |
|  | Self-reported TB contact, or  Living in a household with confirmed TB positive during the survey time | Yes |
| HIV status | Reported not on ART,  Tested negative for HIV currently, and  Self reported as HIV negative when test not available | Negative |
|  | Self reported currently ART use,  Tested positive for HIV currently, or  Self reported as HIV positive when test not available | Positive |

**Table S6:** Distribution of predictors by study for the training data and the test data, unimputed dataset

| **Variable** | **Category** | **Total**  **(N = 169,813)** | | | **South Africa PS**  **(N = 34,573)** | | **TREATS TB**  **(N = 35,315)** | | **ZAMSTAR**  **(N = 54,568)** | | **Zambia PS**  **(N = 45,357)** | |
| --- | --- | --- | --- | --- | --- | --- | --- | --- | --- | --- | --- | --- |
|  |  | **Train (n=135,854)** | **Test (n=33,959)** | **P-value** | **Train (n=27,659)** | **Test (n=6,914)** | **Train (n=28,253)** | **Test (n=7,062)** | **Train (n=43,655)** | **Test (n=10,913)** | **Train** | **Test** |
| Age (years) | Median (IQR) | 31 (22-45) | 31 (22-45) | 0.624^*^ | 37 (25-54) | 37 (25-54 | 29 (21-41) | 29 (21-41) | 29 (23-41) | 29 (23-40) | 32 (22-47) | 32 (22-47) |
|  | *Missing* | 286 (0.2%) | 58 (0.2%) |  | 0 (0.0%) | 0 (0.0%) | 0 (0.0%) | 0 (0.0%) | 286 (0.2%) | 58 (0.2%) | 0 (0.0%) | 0 (0.0%) |
| Sex | Male | 51,386 (37.8%) | 12,753 (37.6%) | 0.358 | 10,634 (38.4%) | 2,585 (37.4%) | 9,967 (35.3%) | 2,424 (34.3%) | 15,489 (35.5%) | 3,904 (35.8%) | 15,296 (42.2%) | 3,840 (42.3%) |
|  | Female | 84,468 (62.2%) | 21,206 (62.4%) |  | 17,025 (61.6%) | 4,329 (62.6%) | 18,286 (64.7%) | 4,638 (65.7%) | 28,166 (64.5%) | 7,009 (64.2%) | 20,991 (57.9%) | 5,230 (57.7%) |
|  | *Missing* | 0 (0.0%) | 0 (0.0%) |  | 0 (0.0%) | 0 (0.0%) | 0 (0.0%) | 0 (0.0%) | 0 (0.0%) | 0 (0.0%) | 0 (0.0%) | 0 (0.0%) |
| Marital status | Single | 47,562 (35.0%) | 11,986 (35.3%) | 0.195 | 0 (0.0%) | 0 (0.0%) | 15,933 (56.4%) | 4,024 (57.0%) | 21,414 (49.1%) | 5,365 (49.2%) | 10,215 (28.2%) | 2,597 (28.6%) |
|  | Married | 47,472 (34.9%) | 11,897 (35.0%) |  | 0 (0.0%) | 0 (0.0%) | 9,168 (32.4) | 2,323 (32.9%) | 17,787 (40.7%) | 4,427 (40.6%) | 20,517 (56.5%) | 5,147 (56.8%) |
|  | Separated/  divorced/widowed | 13,161 (9.7%) | 3,162 (9.3%) |  | 0 (0.0%) | 0 (0.0%) | 3,152 (11.2%) | 715 (10.1%) | 4,454 (10.2%) | 1,121 (10.3%) | 5,555 (15.3%) | 1,326 (14.6%) |
|  | *Missing* | 27,659 (20.4%) | 6,914 (20.4%) |  | 27,659 (100%) | 6,914 (100%) | 0 (0.0%) | 0 (0.0%) | 0 (0.0%) | 0 (0.0%) | 0 (0.0%) | 0 (0.0%) |
| Education | None | 8,850 (6.5%) | 2,130 (6.3%) | 0.110 | 2,509 (9.1%) | 618 (8.9%) | 911 (3.2%) | 211 (3.0%) | 2,046 (4.7%) | 490(4.5%) | 3,384 (9.3%) | 811 (8.9%) |
|  | Primary/ Secondary | 119,765 (88.2%) | 29,941 (88.2%) |  | 23,480 (84.9%) | 5,804 (84.0%) | 26,158 (92.6%) | 6,549 (92.7%) | 38,687 (88.6%) | 9,669 (88.6%) | 31,440 (86.6%) | 7,919 (87.3%) |
|  | Higher | 7,192 (5.3%) | 1,872 (5.5%) |  | 1,646 (6.0%) | 484 (7.0%) | 1,184 (4.2%) | 302 (4.3%) | 2,922 (6.7%) | 754 (6.9%) | 1,440 (4.0%) | 332 (3.7%) |
|  | *Missing* | 47 (0.0%) | 16 (0.1%) |  | 24 (0.1%) | 8 (0.1%) | 0 (0.0%) | 0 (0.0%) | 0 (0.0%) | 0 (0.0%) | 23 (0.1%) | 8 (0.1%) |
| Occupation | Employed full time | 25,814 (19.0%) | 6,418 (18.9%) | 0.858 | 4,352 (15.7%) | 1,169 (16.9%) | 2,835 (10.0%) | 662 (9.4%) | 15,368 (35.2%) | 3,808 (34.9%) | 3,259 (9.0%) | 779 (8.6%) |
|  | Part-time/informal | 30,492 (22.4%) | 7,599 (22.4%) |  | 1,559 (5.6%) | 348 (5.0%) | 5,321 (18.8%) | 1,350 (19.1%) | 7,240 (16.6%) | 1,788 (16.4%) | 16,372 (45.1%) | 4,113 (45.4%) |
|  | Unemployed | 59,390 (43.7%) | 14,918 (43.9%) |  | 17,580 (63.6%) | 4,394 (63.6%) | 15,991 (56.6%) | 3,976 (56.3%) | 14,647 (33.6%) | 3,727 (34.2%) | 11,172 (30.8%) | 2,821 (31.1%) |
|  | Student | 19,188 (14.1%) | 4,767 (14.0%) |  | 3,996 (14.4%) | 954 (13.8%) | 3,673 (13.0%) | 965 (13.7%) | 6,400 (14.7%) | 1,590 (14.6%) | 5,119 (14.1%) | 1,258 (31.1%) |
|  | *Missing* | 970 (0.7%) | 257 (0.8%) |  | 172 (0.6%) | 49 (0.7%) | 433 (1.5%) | 109 (1.5%) | 0 (0.0%) | 0 (0.0%) | 365 (1.0%) | 99 (1.1%) |
| Smoking status | Current smoking | 17,611 (13.0%) | 4,415 (13.0%) | 0.979 | 7,440 (26.9%) | 1,808 (26.2%) | 6,526 (23.1%) | 1,660 (23.5%) | 3,645 (8.4%) | 947 (8.7%) | 0 (0.0%) | 0 (0.0%) |
|  | Not smoking | 81,899 (60.3%) | 20,455 (60.2%) |  | 20,163 (72.9%) | 5,089 (73.6%) | 21,726 (76.9%) | 5,400 (76.5%) | 40,010 (91.6%) | 9,966 (91.3%) | 0 (0.0%) | 0 (0.0%) |
|  | *Missing* | 36,344 (26.8%) | 9,089 (26.8%) |  | 56 (0.2%) | 17 (0.3%) | 1 (0.0%) | 2 (0.0%) | 0 (0.0%) | 0 (0.0%) | 36,287 (100%) | 9,070 (100%) |
| Smoke intensity | Not smoking | 81,899 (60.3%) | 20,455 (60.2%) | 0.922 | 20,163 (72.9%) | 5,089 (73.6%) | 21,726 (76.9%) | 5,400 (76.5%) | 40,010 (91.7) | 9,966 (91.3%) | 0 (0.0%) | 0 (0.0%) |
|  | Low | 1,972 (1.4%) | 510 (1.5%) |  | 385 (1.4%) | 107 (1.6%) | 1,244 (4.4%) | 303 (4.3%) | 343 (0.8%) | 100 (0.9%) | 0 (0.0%) | 0 (0.0%) |
|  | High | 14,821 (10.9%) | 3,700 (10.9%) |  | 6,990 (25.3%) | 1,686 (24.4%) | 5,282 (18.7%) | 1,357 (19.2%) | 2,549 (5.8%) | 657 (6.0%) | 0 (0.0%) | 0 (0.0%) |
|  | *Missing* | 37,162 (27.4%) | 9,294 (27.4%) |  | 121 (0.4%) | 32 (0.5%) | 1 (0.0%) | 2 (0.0%) | 753 (1.7%) | 190 (1.7%) | 36,287 (100%) | 9,070 (100%) |
| Drinking status | Not drinking | 64,619 (47.6%) | 16,168 (47.6%) | 0.987 | 18,177 (65.7%) | 4,599 (66.5%) | 17,899 (63.4%) | 4,498 (63.7%) | 28,543 (65.4%) | 7,071 (64.8%) | 0 (0.0%) | 0 (0.0%) |
|  | Currently drinking | 34,819 (25.6%) | 8,692 (25.0%) |  | 9,429 (34.1%) | 2,302 (33.3%) | 10,278 (36.4%) | 2,548 (36.1%) | 15,112 (34.6%) | 3,842 (35.2%) | 0 (0.0%) | 0 (0.0%) |
|  | *Missing* | 36,416 (26.8%) | 9,099 (26.8%) |  | 53 (0.2%) | 13 (0.2%) | 76 (0.3%) | 16 (0.2%) | 0 (0.0%) | 0 (0.0%) | 36,287 (100%) | 9,070 (100%) |
| Drink intensity | Not drinking | 64,619 (47.6%) | 16,168 (47.6%) | 0.999 | 18,177 (65.7%) | 4,599 (66.5%) | 17,899 (63.4%) | 4,498 (63.7%) | 28,543 (65.4%) | 7,071 (64.8%) | 0 (0.0%) | 0 (0.0%) |
|  | Occasional | 31,998 (23.6%) | 7,986 (23.5%) |  | 8,781 (31.8%) | 2,141 (31.0%) | 9,625 (34.1%) | 2,377 (33.7%) | 13,592 (31.1%) | 3,468 (31.8%) | 0 (0.0%) | 0 (0.0%) |
|  | Heavy | 2,815 (2.1%) | 705 (2.1%) |  | 642 (2.3%) | 160 (2.3%) | 653 (2.3%) | 171 (2.4%) | 1,520 (3.5%) | 374 (3.4%) | 0 (0.0%) | 0 (0.0%) |
|  | *Missing* | 36,422 (26.8%) | 9,100 (26.8%) |  | 59 (0.2%) | 14 (0.2%) | 76 (0.3%) | 16 (0.2%) | 0 (0.0%) | 0 (0.0%) | 36,287 (100%) | 9,070 (100%) |
| Household size | Median (IQR) | 4 (3-6) | 4 (3-6) | 0.128^*^ | 4 (3-6) | 4 (3-6) | NA | NA | 5 (3-7) | 5 (3-7) | NA | NA |
|  | Missing | 64,700 (47.6%) | 16,171 (47.6%) |  | 0 (0.0%) | 0 (0.0%) | 7,062 (100%) | 28,253 (100%) | 160(0.4%) | 39 (0.4%) | 36,287 (100%) | 9,070 (100%) |
| House type | Single unit/brick | 54,792 (40.3%) | 13,594 (40.0%) | 0.350 | 0 (0.0%) | 0 (0.0%) | 13,751 (48.7%) | 3,441 (48.7%) | 23,467 (53.8%) | 5,769 (52.9%) | 17,574 (48.4%) | 4,384 (48.3%) |
|  | Cluster/multi-unit/mudadadas | 14,667 (10.8%) | 3,791 (11.2%) |  | 0 (0.0%) | 0 (0.0%) | 7,086 (25.1%) | 1,801 (25.5%) | 6,471 (14.8%) | 1,705 (15.6%) | 1,110 (3.1%) | 285 (3.1%) |
|  | Flat in block of flats | 8,401 (6.2%) | 2,039 (6.0%) |  | 0 (0.0%) | 0 (0.0%) | 3,147 (11.1%) | 762 (10.8%) | 4,041 (9.3%) | 987 (9.0%) | 1,213 (3.3%) | 290 (3.2%) |
|  | Traditional hut/structure | 18,566 (13.7%) | 4,643 (13.7%) |  | 0 (0.0%) | 0 (0.0%) | 122 (0.4%) | 30 (0.4%) | 6,806 (15.6%) | 1,693 (15.5%) | 11,638 (32.1%) | 2,920 (32.2%) |
|  | Servant quarters or informal dwelling | 5,399 (4.0%) | 1,364 (4.0%) |  | 0 (0.0%) | 0 (0.0%) | 4,109 (14.5%) | 1,020 (14.4%) | 1,169 (2.7%) | 314 (2.9%) | 121 (0.3%) | 30 (0.3%) |
|  | *Missing* | 34,029 (25.1%) | 8,528 (25.1%) |  | 27,659 (100%) | 6,914 (100%) | 38 (0.1%) | 8 (0.1%) | 1,701 (3.9%) | 445 (4.1%) | 4,631 (12.8%) | 1,161 (12.8%) |
| House floor | High quality | 21,945 (16.2%) | 5,544 (16.3%) | 0.777 | 0 (0.0%) | 0 (0.0%) | 8,543 (30.2%) | 2,130 (30.2%) | 13,024 (29.8%) | 3,299 (30.2%) | 378 (1.0%) | 115 (1.3%) |
|  | Medium quality | 56,805 (41.8%) | 14,174 (41.7%) |  | 0 (0.0%) | 0 (0.0%) | 18,316 (64.8%) | 4,591 (65.0%) | 22,722 (52.1%) | 5,648 (51.8%) | 15,767 (43.5%) | 3,935 (43.4%) |
|  | Low quality | 22,280 (16.4%) | 5,510 (16.2%) |  | 0 (0.0%) | 0 (0.0%) | 1,343 (4.8%) | 316 (4.5%) | 6,053 (13.9%) | 1,494 (13.7%) | 14,884 (41.0%) | 3,700 (40.8%) |
|  | *Missing* | 34,824 (25.6%) | 8,731 (25.7%) |  | 27,659 (100%) | 6,914 (100%) | 51 (0.2%) | 25 (0.4%) | 1,856 (4.2%) | 472 (4.3%) | 5,258 (14.5%) | 1,320 (14.6%) |
| Water source | Direct household access | 38,748 (28.5%) | 9,748 (28.7%) | 0.859 | 13,617 (49.2%) | 3,410 (49.3%) | 9,292 (32.9%) | 2,366 (33.5%) | 14,547 (33.3%) | 3,651 (33.5%) | 1,263 (3.5%) | 321 (3.5%) |
|  | On premise | 31,201 (23.0%) | 7,772 (22.9%) |  | 7,144 (25.8%) | 1,835 (26.5%) | 10,148 (35.9%) | 2,438 (34.5%) | 11,633 (26.7) | 2,929 (26.8%) | 2,276 (6.3%) | 570 (6.3%) |
|  | External | 60,370 (44.4%) | 15,034 (44.3%) |  | 6,278 (22.7%) | 1,511 (21.9%) | 8,793 (31.1%) | 2,252 (31.9%) | 17,253 (39.5%) | 4,266 (39.1%) | 28,046 (77.3%) | 7,005 (77.2%) |
|  | *Missing* | 5,564 (4.1%) | 1,405 (4.1%) |  | 620 (2.2%) | 158 (2.3%) | 20 (0.1%) | 6 (0.1%) | 222 (0.5%) | 67 (0.6%) | 4,702 (13.0%) | 1,174 (12.9%) |
| Cook fuel | Clean | 68,776 (50.6%) | 17,298 (50.9%) | 0.580 | 24,693 (89.3%) | 6,171 (89.3%) | 13,564 (48.0%) | 3,400 (48.2%) | 26,788 (61.4%) | 6,730 (61.7%) | 3,731 (10.3%) | 997 (11.0%) |
|  | Unclean | 61,873 (45.5%) | 15,362 (45.2%) |  | 2,444 (8.8%) | 618 (8.9%) | 14,661 (51.9%) | 3,658 (51.8%) | 16,846 (38.6%) | 4,176 (38.3%) | 27,922 (77.0%) | 6,910 (76.2%) |
|  | *Missing* | 5,205 (3.8%) | 1,299 (3.8%) |  | 522 (1.9%) | 125 (1.8%) | 28 (0.1%) | 4 (0.1%) | 21 (0.1%) | 7 (0.1%) | 4,634 (12.8%) | 1,163 (12.8%) |
| Cough | No | 116,421 (85.7%) | 29,037 (85.5%) | 0.373 | 25,328 (91.6%) | 6,340 (91.7%) | 25,416 (90.0%) | 6,326 (89.6%) | 38,463 (88.1%) | 9,574 (87.7%) | 27,214 (75.0%) | 6,797 (74.9%) |
|  | Yes | 19,433 (14.3%) | 4,922 (14.5%) |  | 2,331 (8.4%) | 574 (8.3%) | 2,837 (10.0%) | 736 (10.4%) | 5,192 (11.9%) | 1,339 (12.3%) | 9,073 (25.0%) | 2,273 (25.1%) |
|  | *Missing* | 0 (0.0%) | 0 (0.0%) |  | 0 (0.0%) | 0 (0.0%) | 0 (0.0%) | 0 (0.0%) | 0 (0.0%) | 0 (0.0%) | 0 (0.0%) | 0 (0.0%) |
| Cough duration | No symptom | 116,421 (85.7%) | 29,037 (85.5%) | 0.762 | 25,328 (91.6%) | 6,340 (91.7%) | 25,416 (90.0%) | 6,326 (89.6%) | 38,463 (88.1%) | 9,574 (87.7%) | 27,214 (75.0%) | 6,797 (74.9%) |
|  | <2 weeks | 13,184 (9.7%) | 3,355 (9.9%) |  | 1,094 (4.0%) | 270 (3.9%) | 2,118 (7.5%) | 549 (7.8%) | 3,223 (7.4%) | 817 (7.5%) | 6,749 (18.6%) | 1,719 (19.0%) |
|  | >=2 weeks | 6,069 (4.5%) | 1,525 (4.5%) |  | 1,179 (4.3%) | 292 (4.2%) | 719 (2.5%) | 187 (2.7%) | 1,847 (4.2%) | 492 (4.5%) | 2,324 (6.4%) | 554 (6.1%) |
|  | *Missing* | 180 (0.1%) | 42 (0.1%) |  | 58 (0.2%) | 12 (0.2%) | 0 (0.0%) | 0 (0.0%) | 122 (0.3%) | 30 (0.3%) | 0 (0.0%) | 0 (0.0%) |
| Fever | No | 122,754 (90.4%) | 30,685 (90.4%) | 0.993 | 26,512 (95.9%) | 6,629 (95.9%) | 27,236 (96.4%) | 6,793 (96.2%) | 38,593 (88.4%) | 9,617 (88.1%) | 30,413 (83.8%) | 7,646 (84.3%) |
|  | Yes | 13,100(9.6%) | 3,274 (9.6%) |  | 1,147 (4.2%) | 285 (4.1%) | 1,017 (3.6%) | 269 (3.8%) | 5,062 (11.6%) | 1,296 (11.9%) | 5,874 (16.2%) | 1,424 (15.7%) |
|  | *Missing* | 0 (0.0%) | 0 (0.0%) |  | 0 (0.0%) | 0 (0.0%) | 0 (0.0%) | 0 (0.0%) | 0 (0.0%) | 0 (0.0%) | 0 (0.0%) | 0 (0.0%) |
| Fever duration | No symptom | 122,754 (90.4%) | 30,685 (90.4%) | 0.843 | 26,512 (95.9%) | 6,629 (95.9%) | 27,236 (96.4%) | 6,793 (96.2%) | 38,593 (88.4%) | 9,617 (88.1%) | 30,413 (83.8%) | 7,646 (84.3%) |
|  | <2 weeks | 6,061 (4.5%) | 1,490 (4.4%) |  | 602(2.2%) | 147 (2.1%) | 807 (2.9%) | 214 (3.0%) | 0 (0.0%) | 0 (0.0%) | 4,652 (12.8%) | 1,129 (12.5%) |
|  | >=2 weeks | 1,933 (1.4%) | 480 (1.4%) |  | 501 (1.8%) | 130 (1.9%) | 210 (0.7%) | 55 (0.8%) | 0 (0.0%) | 0 (0.0%) | 1,222 (3.4%) | 295 (3.3%) |
|  | *Missing* | 5,106 (3.8%) | 1,304 (3.8%) |  | 44 (0.2%) | 8 (0.1%) | 0 (0.0%) | 0 (0.0%) | 5,062 (11.6%) | 1,296 (11.9%) | 0 (0.0%) | 0 (0.0%) |
| Night sweats | No | 95,543 (70.3%) | 23,816 (70.1%) | 0.701 | 26,000 (94.0%) | 6,515 (94.2%) | 27,089 (95.9%) | 6,769 (95.9%) | 39,058 (89.5%) | 9,729 (89.2%) | 3,396 (9.4%) | 803 (8.9%) |
|  | Yes | 9,305 (6.9%) | 2,363 (7.0%) |  | 1,659 (6.0%) | 399 (5.8%) | 1,164 (4.1%) | 293 (4.2%) | 4,597 (10.5%) | 1,184 (10.9%) | 1,885 (5.2%) | 487 (5.4%) |
|  | *Missing* | 31,006 (22.8%) | 7,780 (22.9%) |  | 0 (0.0%) | 0 (0.0%) | 0 (0.0%) | 0 (0.0%) | 0 (0.0%) | 0 (0.0%) | 31,006 (85.5%) | 7,780 (85.8%) |
| Weight loss | No | 94,844 (69.8%) | 23,561 (69.4%) | 0.08 | 26,413 (95.5%) | 6,585 (95.2%) | 27,034 (95.7%) | 6,735 (95.4%) | 37,925 (86.9%) | 9,389 (86.0%) | 3,472 (9.6%) | 852 (9.4%) |
|  | Yes | 10,004 (7.4%) | 2,618 (7.7%) |  | 1,246 (4.5%) | 329 (4.8%) | 1,219 (4.3%) | 327 (4.6%) | 5,730 (13.1%) | 1,524 (14.0%) | 1,809 (5.0%) | 438 (4.8%) |
|  | *Missing* | 31,006 (22.8%) | 7,780 (22.9%) |  | 0 (0.0%) | 0 (0.0%) | 0 (0.0%) | 0 (0.0%) | 0 (0.0%) | 0 (0.0%) | 31,006 (85.5%) | 7,780 (85.8%) |
| Chest pain | No | 91,355 (67.2%) | 22,840 (67.3%) | 0.998 | 0 (0.0%) | 0 (0.0%) | 26,429 (93.5%) | 6,575 (93.1%) | 39,658 (90.8%) | 9,918 (90.9%) | 25,268 (69.6%) | 6,347 (70.0%) |
|  | Yes | 16,840 (12.4%) | 4,205 (12.4%) |  | 0 (0.0%) | 0 (0.0%) | 1,824 (6.5%) | 487 (6.9%) | 3,997 (9.2%) | 995 (9.1%) | 11,019 (30.4%) | 2,723 (30.0%) |
|  | *Missing* | 27,659 (20.4%) | 6,914 (20.4%) |  | 27,659 (100%) | 6,914 (100%) | 0 (0.0%) | 0 (0.0%) | 0 (0.0%) | 0 (0.0%) | 0 (0.0%) | 0 (0.0%) |
| Chest pain duration | No symptom | 91,355 (67.2%) | 22,840 (67.3%) | 0.9999 | 0 (0.0%) | 0 (0.0%) | 26,429 (93.5%) | 6,575 (93.1%) | 39,658 (90.8%) | 9,918 (90.9%) | 25,268 (69.6%) | 6,347 (70.0%) |
|  | <2 weeks | 8,921 (6.6%) | 2,227 (6.6%) |  | 0 (0.0%) | 0 (0.0%) | 960 (3.4%) | 253 (3.9%) | 0 (0.0%) | 0 (0.0%) | 7,961 (21.9%) | 1,974 (21.8%) |
|  | >=2 weeks | 3,922 (2.9%) | 983 (2.9%) |  | 0 (0.0%) | 0 (0.0%) | 864 (3.1%) | 234 (3.3%) | 0 (0.0%) | 0 (0.0%) | 3,058 (8.4%) | 749 (8.3%) |
|  | *Missing* | 31,656 (23.3%) | 7,909 (23.3%) |  | 27,659 (100%) | 6,914 (100%) | 0 (0.0%) | 0 (0.0%) | 3,997 (9.2%) | 995 (9.1%) | 0 (0.0%) | 0 (0.0%) |
| TB contact | No | 74,194 (54.6%) | 18,554 (54.6%) | 0.532 | 0 (0.0%) | 0 (0.0%) | 26,849 (95.0%) | 6,714 (95.1%) | 42,265 (96.8%) | 10,589 (97.0%) | 5,000 (14.0%) | 1,251 (13.8%) |
|  | Yes | 2,897 (2.1%) | 691 (2.0%) |  | 0 (0.0%) | 0 (0.0%) | 1,049 (3.7%) | 248 (3.5%) | 1,390 (3.2%) | 324 (3.0%) | 458 (1.3%) | 119 (1.3%) |
|  | *Missing* | 58,763 (43.3%) | 14,714 (43.3%) |  | 27,659 (100%) | 6,914 (100%) | 355 (1.3%) | 100 (1.4%) | 0 (0.0%) | 0 (0.0%) | 30,749 (84.7%) | 7,700 (84.9%) |
| Previously treated for TB | No | 95,543 (70.3%) | 23,812 (70.1%) | 0.670 | 25,305 (91.5%) | 6,292 (91.0%) | 25,512 (90.3%) | 6,393 (90.5%) | 39,891 (91.4%) | 9,938 (91.1%) | 4,835 (13.3%) | 1,189 (13.1%) |
|  | Yes | 9,050 (6.7%) | 2,301 (6.8%) |  | 2,263 (8.2%) | 590 (8.5%) | 2,600 (9.2%) | 638 (9.0%) | 3,757 (8.6%) | 973 (8.9%) | 430 (1.2%) | 100 (1.1%) |
|  | *Missing* | 31,261 (23.0%) | 7,846 (23.1%) |  | 91 (0.3%) | 32 (0.5%) | 141 (0.5%) | 31 (0.4%) | 7 (0.0%) | 2 (0.0%) | 31,022 (85.5%) | 7,781 (85.8%) |
| Times treated for TB | Never | 65,196 (48.0%) | 16,230 (47.8%) | 0.478 | 25,305 (91.5%) | 6,292 (91.0%) | 0 (0.0%) | 0 (0.0%) | 39,891 (91.4%) | 9,938 (91.1%) | 0 (0.0%) | 0 (0.0%) |
|  | Once | 5,128 (3.8%) | 1,321 (3.9%) |  | 2,051 (7.4%) | 532 (7.7%) | 0 (0.0%) | 0 (0.0%) | 3,077 (7.1%) | 789 (7.2%) | 0 (0.0%) | 0 (0.0%) |
|  | Twice+ | 868 (0.6%) | 236 (0.7%) |  | 197 (0.7%) | 53 (0.8%) | 0 (0.0%) | 0 (0.0%) | 671 (1.5%) | 183 (1.7%) | 0 (0.0%) | 0 (0.0%) |
|  | *Missing* | 64,662 (47.6%) | 16,172 (47.6%) |  | 106 (0.4%) | 37 (0.5%) | 28,253 (100%) | 7,062 (100%) | 16 (0.0%) | 3 (0.0%) | 36,287 (100%) | 9,070 (100%) |
| HIV | Negative | 83,264 (61.3%) | 20,852 (61.4%) | 0.813 | 17,529 (63.4%) | 4,416 (63.9%) | 16,646 (58.9%) | 4,152 (58.8%) | 26,682 (61.1%) | 6,645 (60.9%) | 22,407 (61.8%) | 5,639 (62.2%) |
|  | Positive | 15,135 (11.1%) | 3,802 (11.2%) |  | 3,566 (12.9%) | 889 (12.9%) | 4,705 (16.7%) | 1,197 (17.0%) | 5,244 (12.0%) | 1,317 (12.1%) | 1,620 (4.5%) | 399 (4.4%) |
|  | *Missing* | 37,455 (27.6%) | 9,305 (27.4%) |  | 6,564 (23.7%) | 1,609 (23.3%) | 6,902 (24.4%) | 1,713 (24.3%) | 11,729 (26.9%) | 2,951 (27.0%) | 12,260 (33.8%) | 3,032 (33.4%) |
| Diabetes | No | 67,514 (49.7%) | 16,876 (49.7%) | 0.838 | 25,828 (93.4%) | 6,479 (93.7%) | 0 (0.0%) | 0 (0.0%) | 41,686 (95.5%) | 10,394 (95.2%) | 0 (0.0%) | 0 (0.0%) |
|  | Yes | 3,395 (2.5%) | 830 (2.4%) |  | 1,426 (5.2%) | 311 (4.5%) | 0 (0.0%) | 0 (0.0%) | 1,969 (4.5%) | 519 (4.8%) | 0 (0.0%) | 0 (0.0%) |
|  | *Missing* | 64,945 (47.8%) | 16,256 (47.9%) |  | 405 (1.5%) | 124 (1.8%) | 28,253 (100%) | 7,062 (100%) | 0 (0.0%) | 0 (0.0%) | 36,287 (100%) | 9,070 (100%) |

^*^P-value from the Mann-Whitney U test, while the rest are p-values based on the chi-square test of association

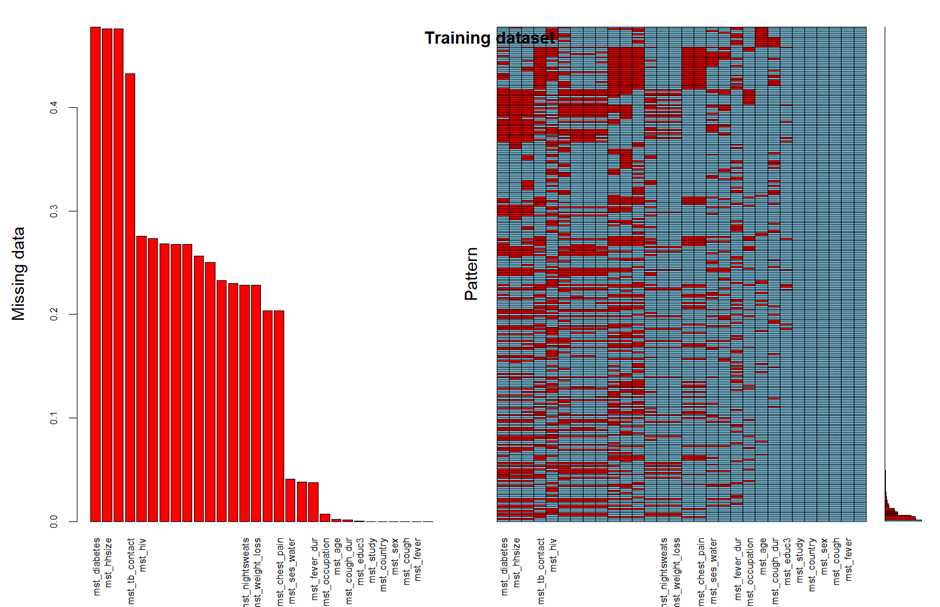

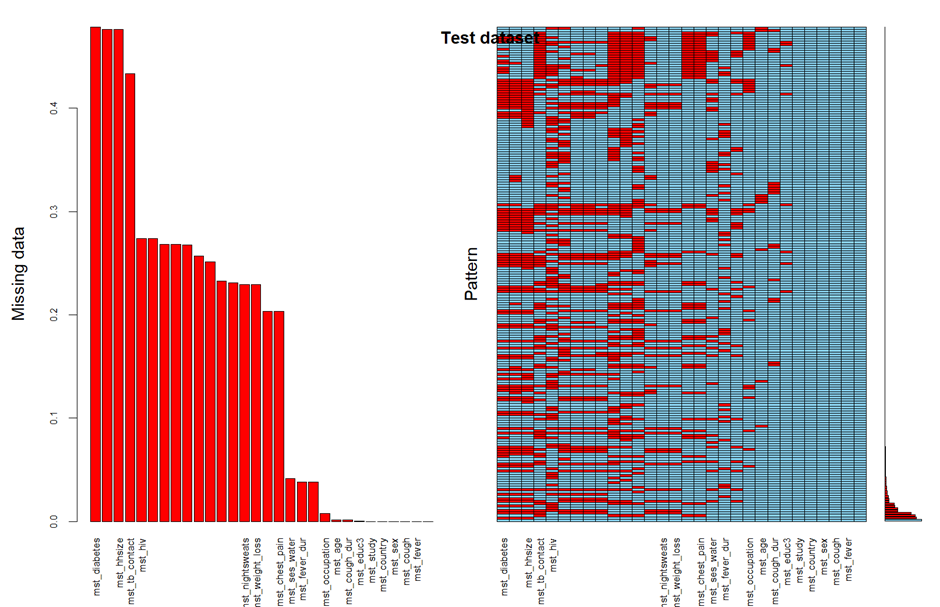

**Figure S1: Visualization of missing data pattern: the training (top) and test (bottom) datasets.** The figures consist of two panels. The left panel displays the proportion of missing data for each variable, where the height of each bar represents the fraction of observations with missing values. Variables are ordered according to the magnitude of missingness, with those exhibiting higher proportions of missing data appearing first. The right panel presents a missing data pattern matrix where each column corresponds to a variable and each row represents a distinct pattern of missingness observed in the dataset. Cells are color-coded to indicate data availability: blue denotes observed values, while red denotes missing values. The frequency of each missing data pattern across observations is displayed by the small bar plot on the far right of the matrix, indicating how often each combination of missing values occurs. The visualization indicated varying proportions of missingness across variables. The missing data matrix showed heterogeneous and non-monotone missing patterns across observations, with no single dominant pattern. Several observations contained complete data across all variables, while others exhibited different combinations of missing values. The presence of multiple missingness patterns and partially observed variables suggests that missingness may plausibly depend on observed covariates rather than unobserved values, supporting the Missing at Random (MAR) assumption underlying multiple imputation using chained equations implemented in the mice.

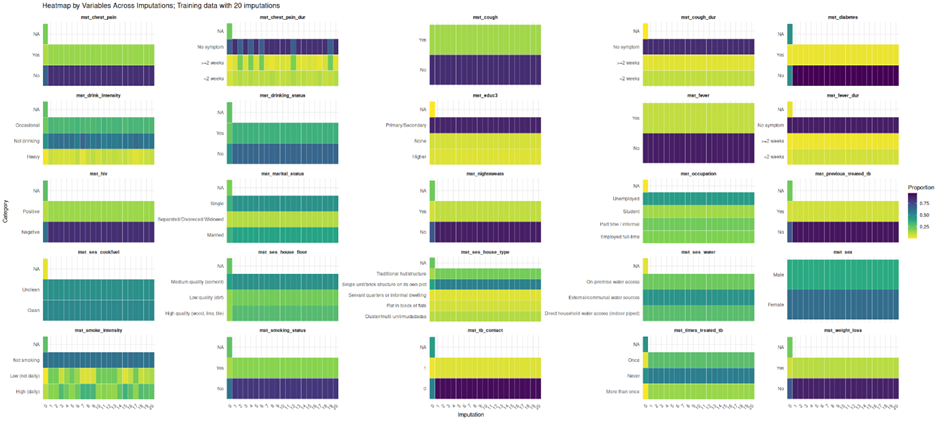

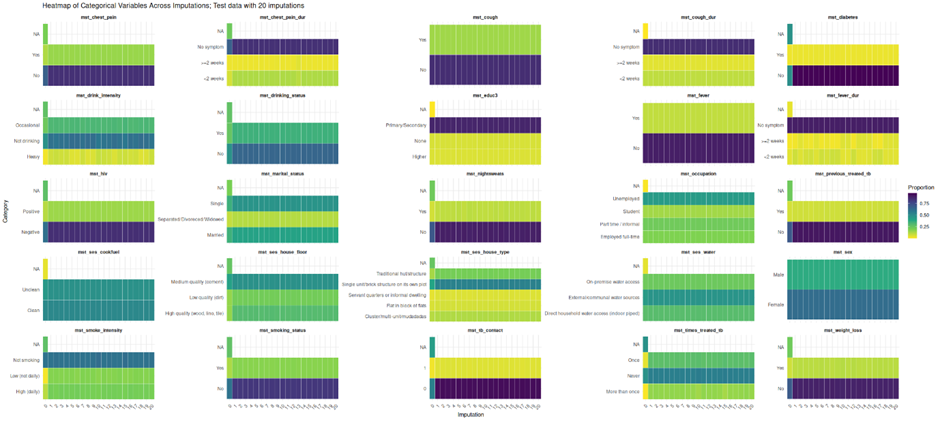

**Figure S2: Comparison of variable distributions across multiple imputations and observed datasets.** Heatmaps illustrate the categorical distributions of features within the training (top) and test (bottom) datasets across **2**0 independent multiple imputation iterations and one **original** dataset. The consistent color patterning across the x-axis indicates that the multiple imputation process maintained stable variable distributions, with the imputed values remaining highly consistent across all iterations and aligning closely with the observed data profile. NA, not available (missing).

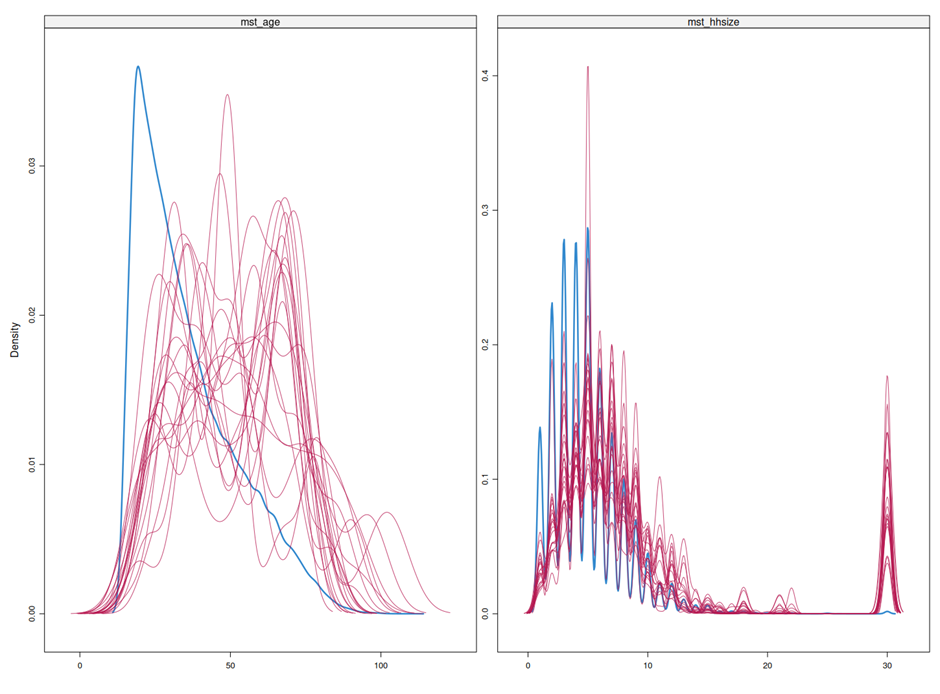

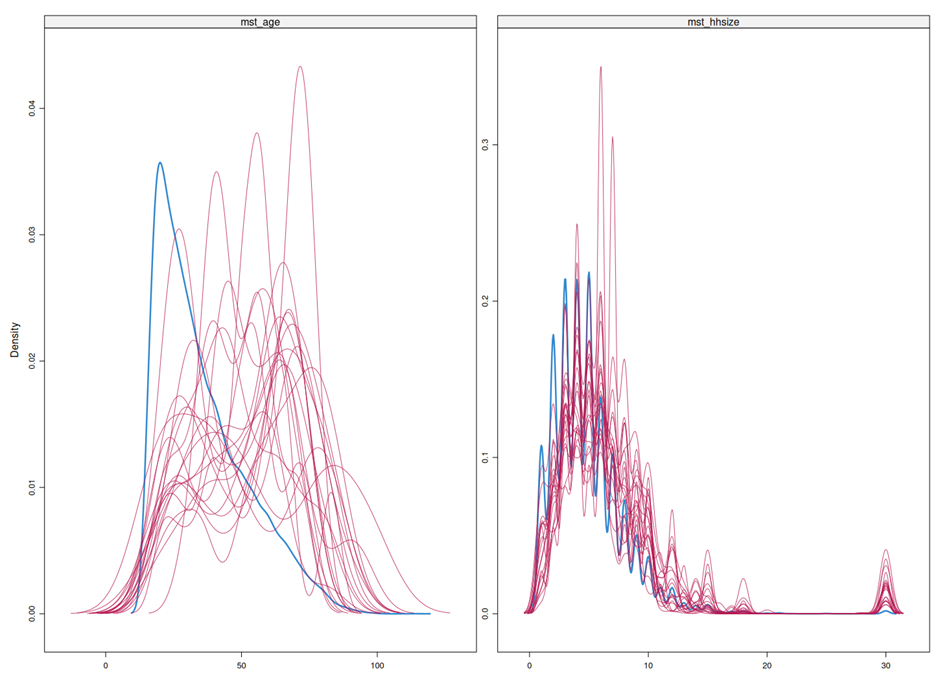

**Figure S3:** Density plots of the training (top) and the test (bottom) datasets across 20 multiple imputations generated using MICE for the variable age and household size. The blue line shows the observed data distribution, while the red lines represent the distributions from each imputed dataset. The overlap between observed and imputed distributions suggests that the imputations reasonably preserve the underlying data structure.

**Table S7.** Summary of hyperparameters tuned in the mTBScreen XGBoost model.

| **Hyperparameter** | **Description** | **Search space** | **Value used** |
| --- | --- | --- | --- |
| n_estimators | The number of trees the model will build. | (250, 500) | 385 |
| max_depth | Maximum level of each tree. Indicates how complex each tree can become. | (20, 30) | 15 |
| gamma | How much improvement is needed to add another split in a tree. | 10 | 5 |
| alpha | L1 regularization. Controls that help prevent the model from becoming too complex and overfitting. | (0.01, 5, 'log-uniform') | 4.99 |
| lambda | L2 regularization. Controls that help prevent the model from becoming too complex and overfitting. | (1, 5, 'log-uniform'), | 5 |
| eta | The learning rate that controls how quickly the model learns from the training data. | (0.001, 0.05, 'log-uniform') | 0.01 |
| subsample | Subsample ratio of the training instance. | (0.75, 1.0, 'uniform') | 0.75 |
| colsample_bytree | Subsample ratio of columns when constructing each tree. | (0.75, 1.0, 'uniform') | 0.75 |
| colsample_bylevel | Subsample ratio of columns for each level. | (0.75, 1.0, 'uniform') | 0.75 |
| max_delta_step | Maximum delta step we allow each tree’s weight estimation to be. | (0, 10) | 0 |

**Note S1.** Performance metrics used to evaluate mTBScreen.

Sensitivity (equivalent to the recall on the positive outcome group), measure the proportion of correctly identified positive instances over all actual positive instances.

$$Sensitivity=\frac{True Positives}{True Positives + False Negatives}$$

Specificity (equivalent to the recall on the negative outcome group), measure the proportion of correctly identified negative instances over all actual negative instances.

$$Specificity=\frac{True Negatives}{True Negatives + False Positives}$$

Positive predictive value (PPV) (equivalent to the precision of the positive outcome group) is the proportion of correctly identified positive instances among all predicted as positive.

$$PPV=\frac{True Positives}{True Positives + False Positives}$$

Negative predictive value (NPV) (equivalent to the precision of the negative outcome group) is the proportion of correctly identified negative instances among all predicted as negative.

$$NPV=\frac{True Negatives}{True Negatives + False Negatives}$$

The Brier score measuring the overall accuracy of probabilistic predictions for a binary outcome Y, defined as the mean squared difference between predicted probabilities and observed outcomes:

$$Brier Score=\frac{1}{n}\sum_{i=1}^{n} {(\hat{p}_{i}-Y_{i})}^{2}$$

Calibration-in-the-large and calibration slope were estimated using logistic calibration regression, fitting a generalized linear model with binomial family:

$$logit\left( P\left( Y=1|logit\left( \hat{p} \right) \right) \right)=\alpha+ \beta*logit(\hat{p})$$

where Y denotes the observed binary outcome and $\hat{p}$. the predicted probability from the model. The calibration intercept ($\alpha$) evaluates systematic over- or underprediction, with $\alpha$ = 0 indicating perfect agreement in baseline risk. The calibration slope ($\beta$) evaluates the spread of predictions, with $\beta$ = 1 indicating ideal calibration; $\beta$ < 1 suggests that estimated risks are too extreme (too high for patients who are at high risk and too low for patients who are at low risk), whereas $\beta$ > 1 indicates compressed predictions (risk estimates are too moderate).

**Note S2.** Net benefit calculation; Decision curve analysis

The net benefit in decision curve analysis for clinical utility was calculated for each threshold (*p_t_*) as:^1^

$$Net Benefit=\frac{True positive count}{n}-\frac{False positive count}{n}*(\frac{p_{t}}{1-p_{t}})$$

In this framework, the two primary clinical harms are accounted. The harm of over-testing (predicted false positives) is explicitly penalized by the formula as the proportion of predicted false positives, weighted by the harm:benefit trade-off ratio $\frac{p_{t}}{1-p_{t}}$. The harm of under-testing (predicted false negatives) is implicitly penalized, as a missed case fails to contribute to the "Benefit" term $\frac{True positive count}{n}$. This concept of computing net benefit could be used for the desired model as well as for other comparator models that lead us to identify range of thresholds where our model has better net clinical benefit.

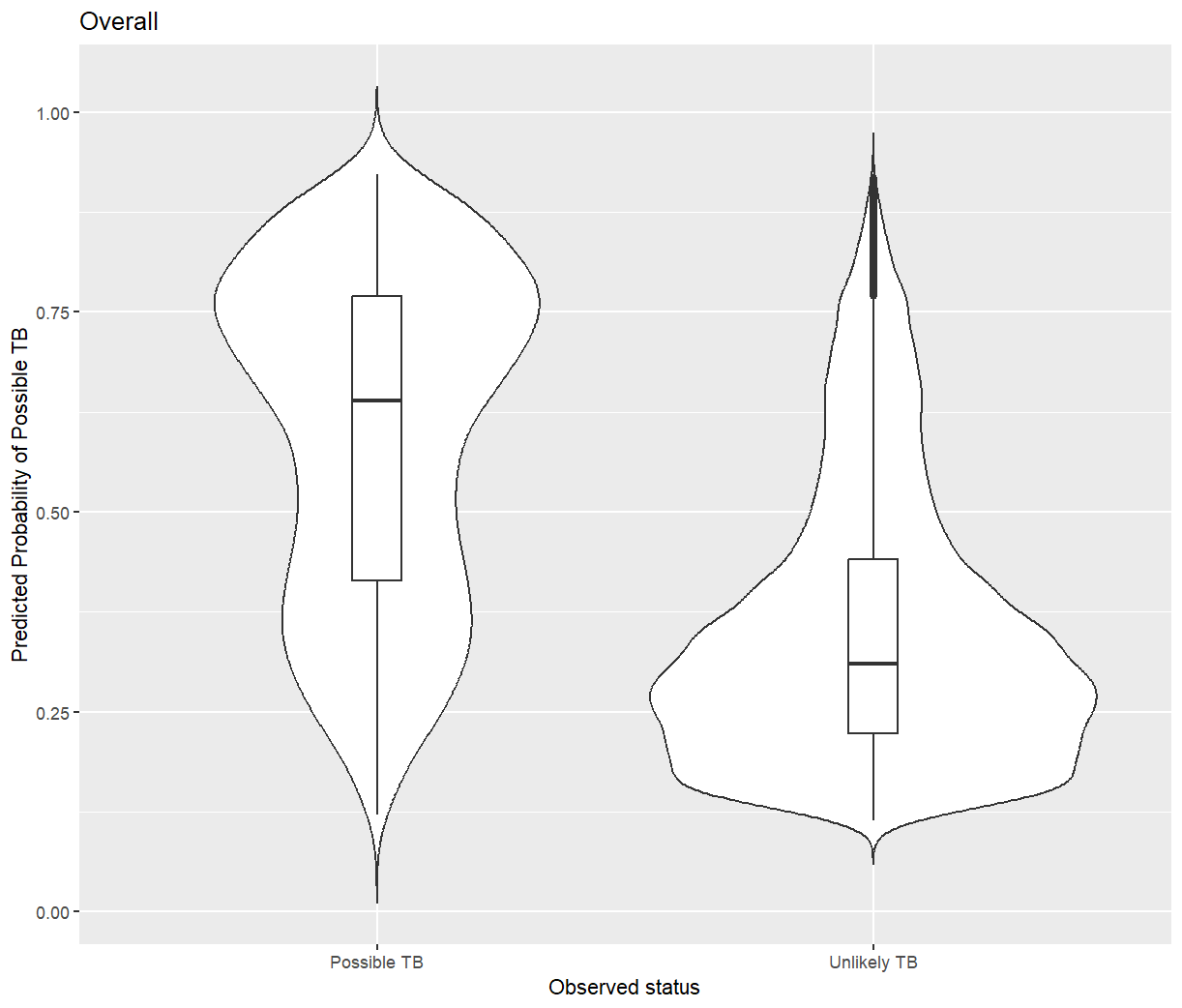

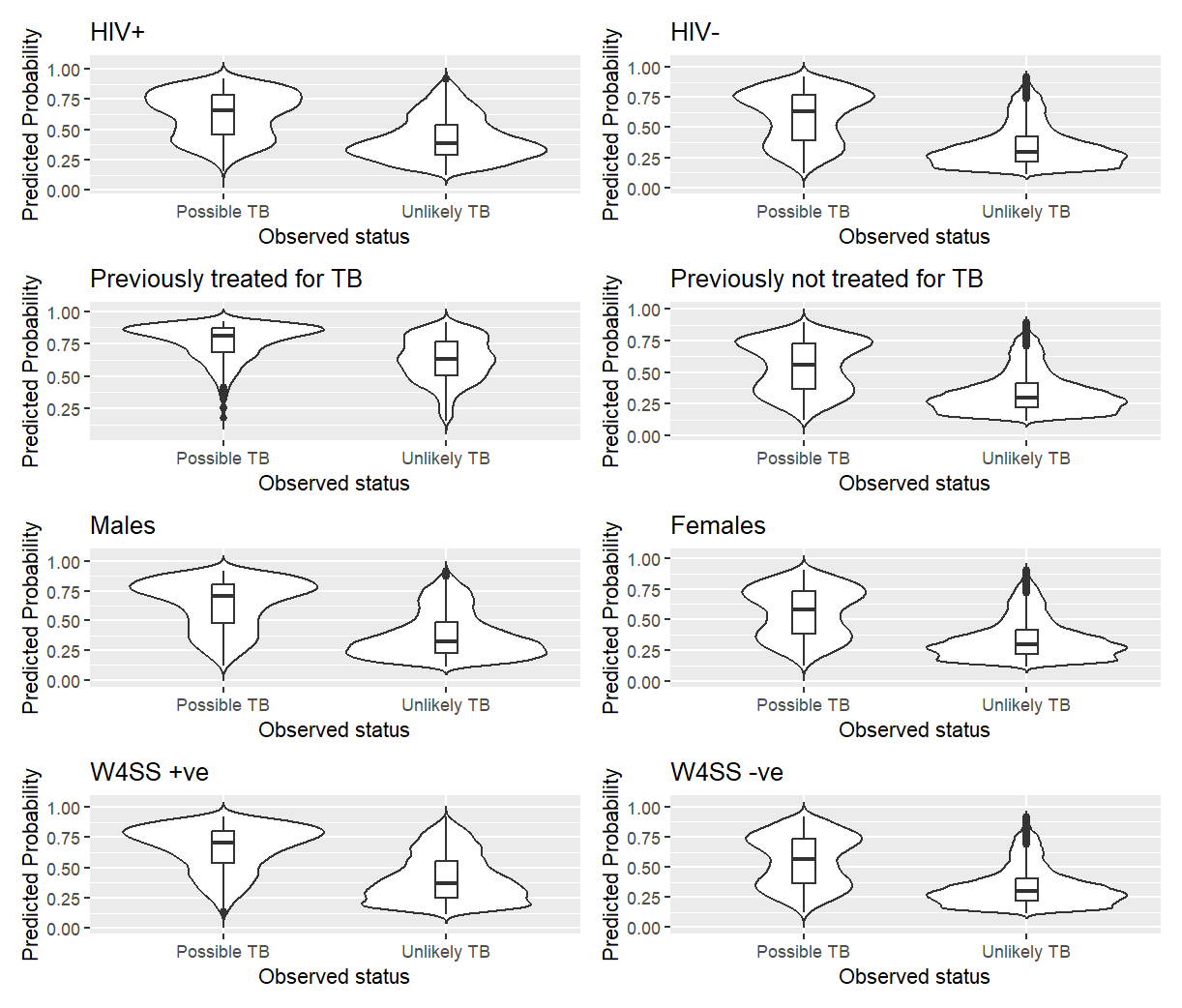

**Figure S4: Predicted probability distributions for 'Possible TB' stratified by observed status and clinical strata.** Violin plots show the density of predicted probabilities for 'Possible TB' for the overall observations (left) and across different variable strata (right). The x-axis shows the prediction distribution by the observed clinical classification (Possible TB vs. Unlikely TB). Internal boxplots indicate the median and interquartile range (IQR). The model demonstrates consistent discriminatory performance across all strata, including HIV status, history of TB treatment, sex, and W4SS results, consistently assigning higher probabilities to individuals with an observed 'Possible TB' status.

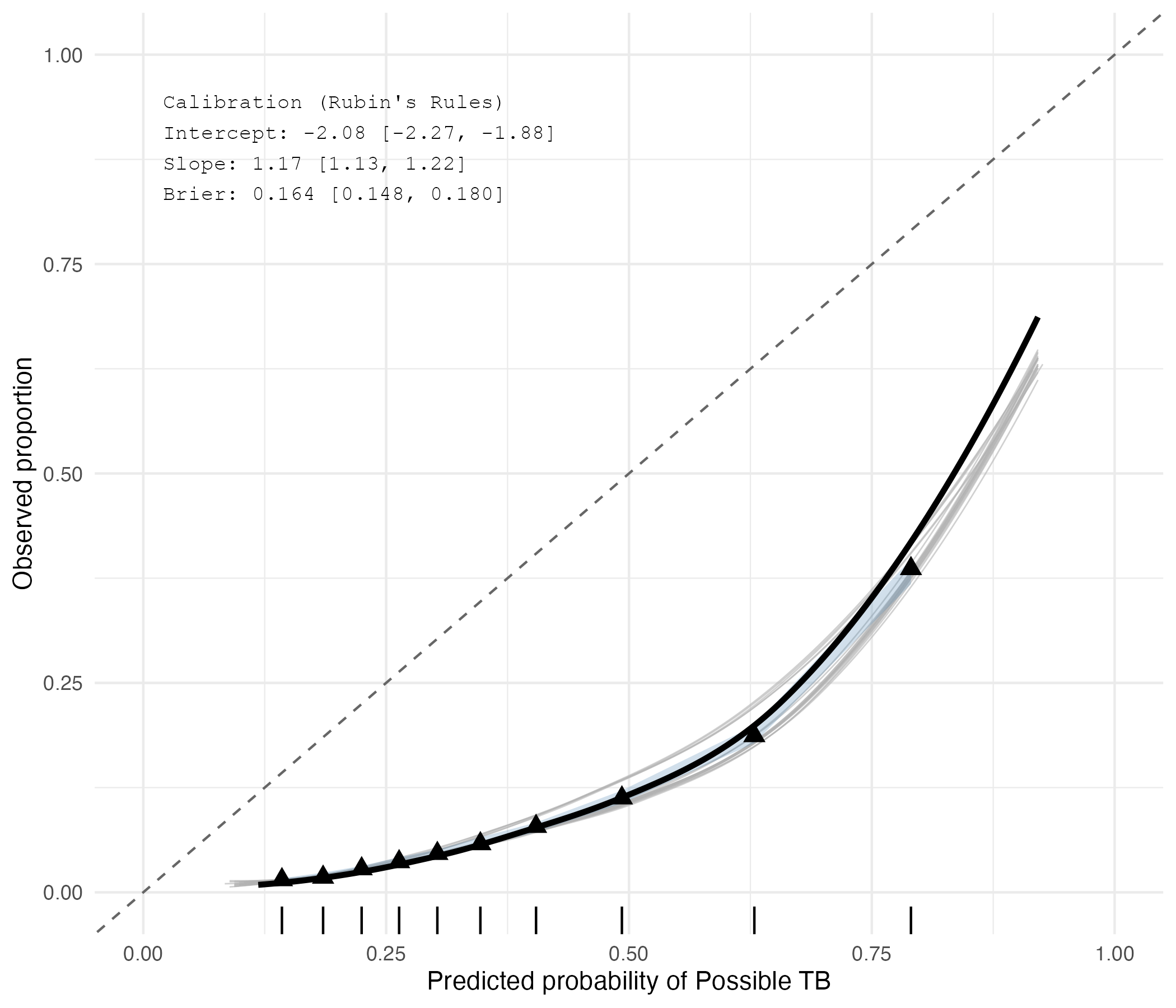
 **Figure S3: Calibration of XGBoost model probabilities via LOESS smoothing and decile analysis.** Predicted probabilities for 'Possible TB' (x-axis) are plotted against observed proportions (y-axis). The light grey lines represent the individual LOESS smoothed curves for each of the 20 imputed datasets, while the bold black line and triangles represent the pooled ensemble predictions and decile bins, respectively. The dashed diagonal represents an ideal calibration (intercept of 0, slope of 1). The position of the smoothed curves below the reference line demonstrates a systematic overestimation of TB risk. This aligns with the pooled calibration metrics computed using Rubin's rules (intercept = -2.08 [95% CI: -2.27, -1.88]; calibration slope = 1.17 [95% CI: 1.13, 1.22]; Brier score = 0.16 [95% CI: 0.15, 0.18]).

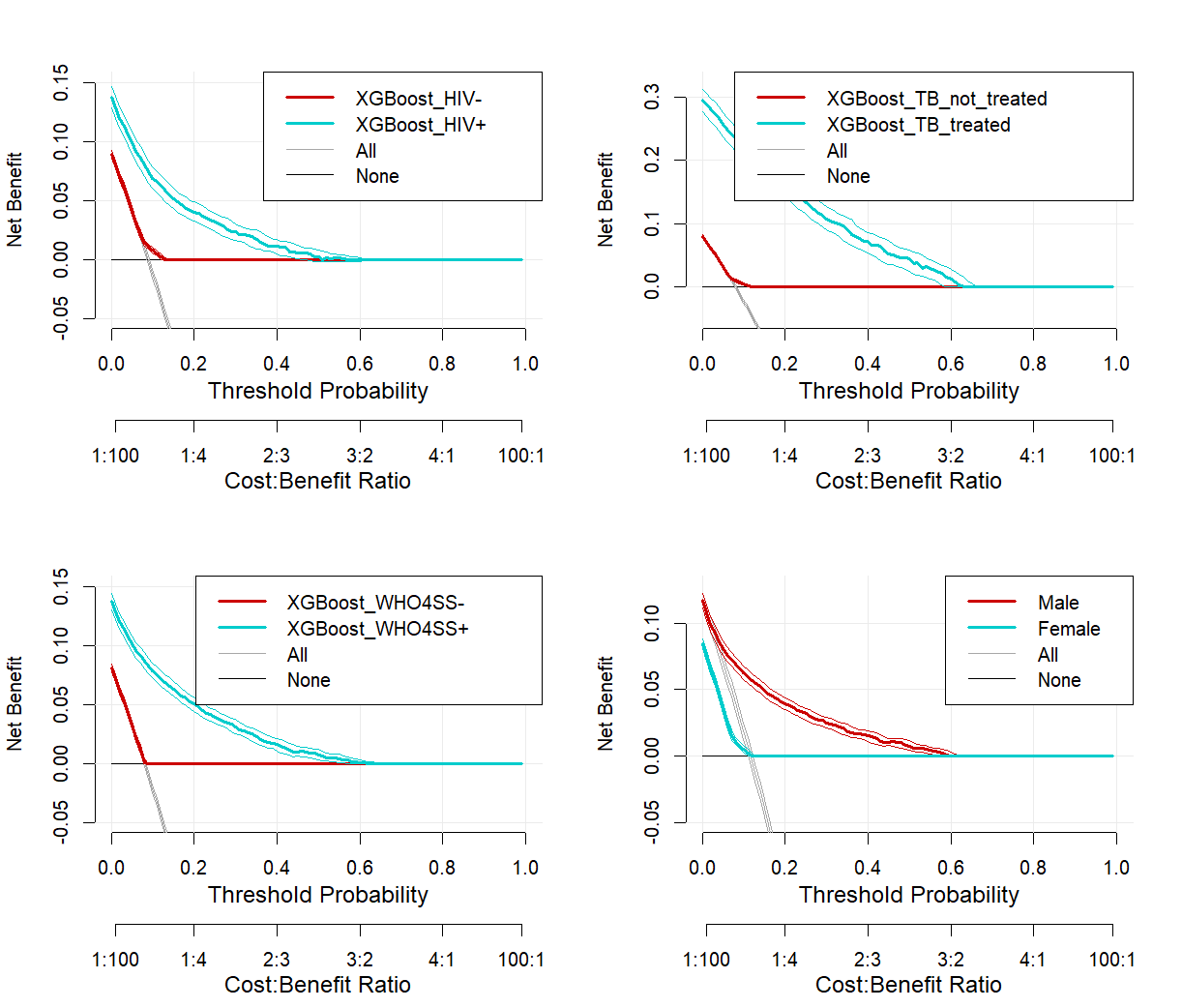

**Figure S4. Decision Curve Analysis (DCA) evaluating the clinical utility of the ML model across clinical subgroups.** The plots illustrate the net clinical benefit (y-axis) of using the XGBoost model compared to "treat-all" (gray line) and "treat-none" (horizontal black line) strategies. The model demonstrates positive net benefit across a wide range of threshold probabilities, with notably higher clinical utility for HIV-positive individuals, those previously treated for TB, symptomatic (W4SS+) individuals, and males. The secondary x-axis displays the corresponding cost-benefit ratios for clinical decision-making.
